# The value of a family history to distinguish monogenic from polygenic amyotrophic lateral sclerosis

**DOI:** 10.64898/2026.09.16.26362994

**Authors:** Paul Beele, Jan H. Veldink, Wouter van Rheenen

**Affiliations:** Department of Neurology, UMC Utrecht Brain Center, University Medical Center Utrecht, Utrecht University, Utrecht 3584 CG, The Netherlands

**Keywords:** familial ALS, sporadic ALS, fALS criteria, simulation framework

## Abstract

Familial recurrence of amyotrophic lateral sclerosis (ALS) or frontotemporal dementia (FTD) is the clinical cornerstone for assuming Mendelian, monogenic inheritance which informs genetic testing, counselling and gene-discovery strategies in standard practice. Polygenic inheritance can lead to familial clustering of ALS, while pathogenic variants with reduced penetrance may present sporadically in small, modern-day pedigrees. As both rare genetic variants and polygenic disease contribute to the occurrence of ALS, the accuracy of a positive family history to predict the underlying genetic architecture remains unknown.

Using growing knowledge of the genetic architecture of ALS, we designed the APECS simulator: a large-scale pedigree simulator using historical demographic parameters, unifying frameworks for monogenic and polygenic disease models. We simulated 1,000,000 three-generation pedigrees and modelled ALS, FTD, and other dementias under both inheritance models. For index patients with ALS, we examined the performance of proposed familial ALS (fALS) definitions to predict the underlying genetic architecture. Simulation results were benchmarked against real-world population estimates, over a wide range of demographic and genetic parameters. We assessed the phenocopy rate for monogenic index patients, defined as relatives with polygenic ALS or a pathogenic allele from a different ancestor. Finally, we illustrated the value of distant relatedness among apparently sporadic, but monogenic ALS patients.

Simulations generated 55.3 million individuals, yielding 5,431 ALS-affected index patients (16.4% monogenic). The proposed criterion for ‘definite fALS’ (≥2 relatives with ALS among relatives within two degrees) had a 31.1% sensitivity, while falsely classifying 10.0% of ‘fALS’ as monogenic. The sensitivity of ‘definite fALS’ was improved to 43.3% when also considering relatives with FTD. Considering any type of dementia raised the false positive rate to 35.9%. In total, 5.3% of monogenic index patients had a phenocopy with ALS in relatives up to the third degree. Of seemingly sporadic monogenic index patients, 83.1% had distant relatives with ALS/FTD up to the fifth degree.

A family history provides limited accuracy for inferring genetic architecture in ALS. Considering unspecified dementias in the family history reduces this accuracy, while adding definite FTD improves accuracy. Relying on traditional fALS criteria risks both missing cases for genetic testing and treatments, and falsely alarming families with polygenic ALS. Given the limited value of a family history and risk of phenocopies, routine genetic testing regardless of family history and systematic approaches for detecting distant relatedness will help to find known and novel genetic causes in ALS.

## Introduction

Amyotrophic lateral sclerosis (ALS) is a fatal neurodegenerative disorder with a complex genetic architecture at the intersect of truly monogenic diseases and polygenic disease traits. It is commonly stated that 10 - 15% of cases are classified as familial ALS (fALS) suggesting a monogenic cause, while most patients are sporadic, reflecting a complex polygenic architecture combined with environmental risk factors.^1^ Consequently, the familial recurrence of ALS or frontotemporal dementia (FTD) is considered instrumental to decide on genetic testing, counselling families on disease risk, and the design of gene-discovery studies in both the clinical and research setting.^2^ This simplified approach overlooks the fact that the utility of a family history for inferring the underlying genetic architecture of ALS/FTD at the individual level remains unknown.

First, there is no consensus on the criteria for a positive family history to fulfil the definition of fALS and which diseases should be considered (e.g. ALS, FTD, and/or unspecified dementia).^1^ Reported fALS criteria range from “at least one relative with ALS no matter how distant” to “at least two first-degree relatives with ALS”.^3,4^ Second, the distinction between familial and sporadic ALS does not always reflect underlying genetic architecture. Complex trait genetics theory dictates that familial clustering can still occur in polygenic disorders.^5^ Conversely, even highly penetrant, monogenic ALS mutations can manifest as apparently sporadic cases in small modern-day families,^6^ as fertility rates continue to decline.^7^ The distinction between familial and sporadic ALS is further obscured by recent studies that show that several pathogenic ALS mutations are less penetrant than previously assumed.^8,9^

In practice, it is challenging to accurately evaluate predictive values of different definitions for fALS based on family histories and genetic testing results. Reliable interpretation of the family history is limited by patients’ reporting accuracy and clinicians’ biased assessments.^3^ Family histories may be recorded more thoroughly when monogenic inheritance is suspected. Especially in a late-onset disease as ALS, the patient-reported information about (distant) relatives is often incomplete. Moreover, to confirm or rule out a monogenic disease, a perfect genetic test is required. In practice, however, not all patients have undergone comprehensive genetic testing, and the pathogenicity of detected variants frequently remains uncertain.^8^ Furthermore, the list of identified genetic causes for ALS is not complete yet.^10^

We can overcome these limitations by leveraging the increasing knowledge on genetic causes and heritability of ALS in population simulations. In this study, we simulated large-scale pedigrees using a newly developed simulation framework called APECS (*ALS Pedigree simulations under a Complex and Simple disease model*). This pedigree simulator uses empirically derived epidemiological, general population and genetic parameters for ALS, FTD, and other dementias, incorporating both monogenic and polygenic inheritance models. We then evaluated the performance of varying fALS definitions to estimate the probability of monogenic inheritance. We compared simulated disease occurrence to real-world population estimates as benchmarks and conducted extensive sensitivity analyses across parameter ranges to assess the robustness and accuracy of fALS classification criteria. Finally, we quantified the expected occurrence of polygenic ALS among relatives of monogenic index patients (i.e. “phenocopies”) and the value of extending family histories to distant relatives to identify apparently sporadic, monogenic cases.

## Materials and methods

### Pedigree simulation

#### Demographic parameters

Pedigrees were simulated to model multigenerational ALS, FTD, and dementia families (Fig. 1). The simulation parameters based on literature estimates are listed in Supplementary Table 1.^7–9,11–29^ Supplementary File 1A provides a detailed outline and illustration of the simulation process.

**Figure 1:**
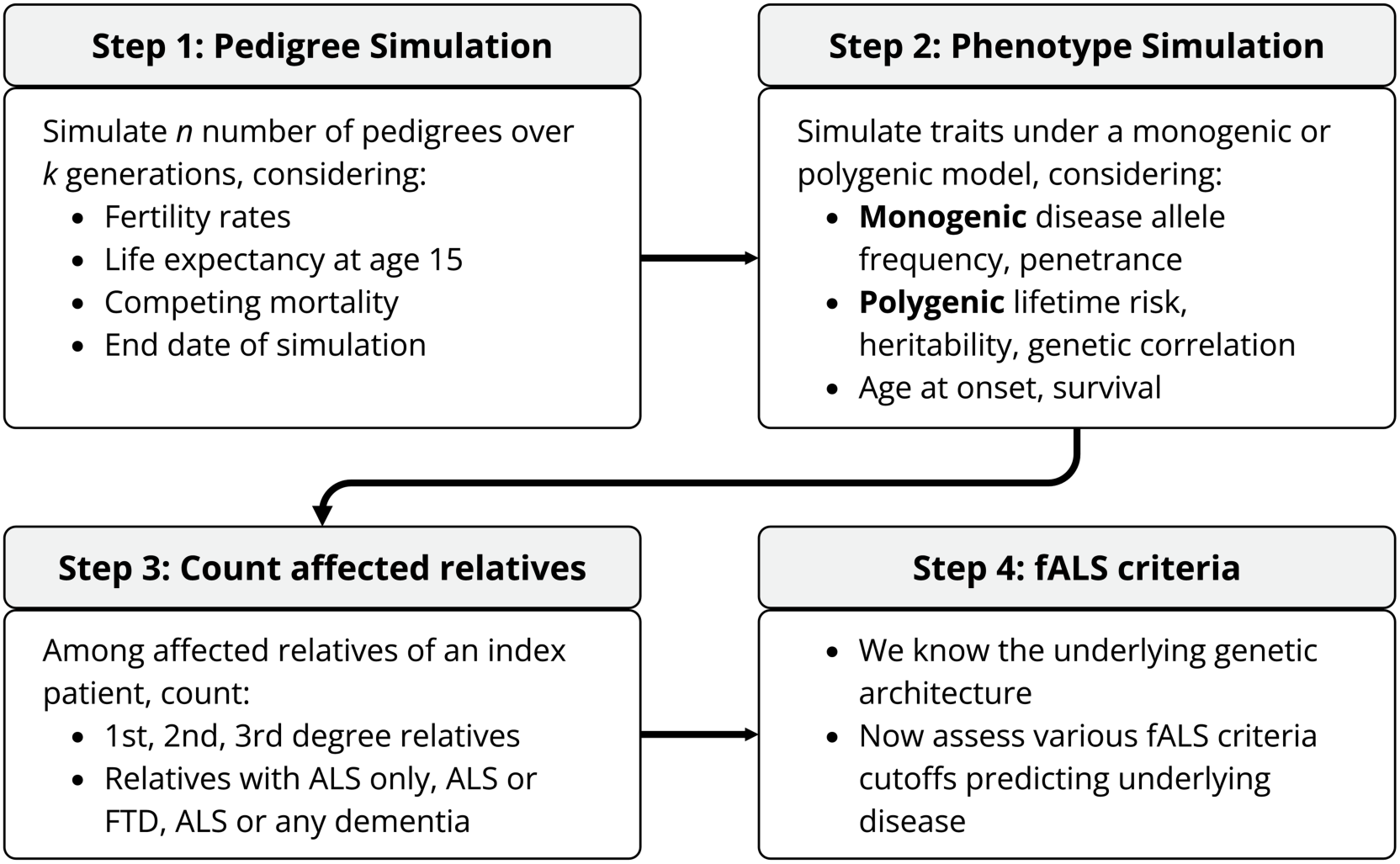
Overview of simulation workflow. Pedigrees were simulated based on historical demographic parameters. Phenotypic traits were simulated under a monogenic and polygenic disease model. For each index patient, affected relatives up to the third degree were counted. The number of affected relatives was then used as a predictor for underlying genetic architecture.

In summary, we modelled historically accurate family structures for three-generation core pedigrees with in-law branches. The index patient generation’s year of birth was sampled between 1940 and 2000 to reflect currently diagnosed ALS patients. For each individual in the pedigree, we sampled life expectancies from historical data to account for competing mortality (Fig. 2A-D). We used historical life expectancies at age 15 to account for childhood mortality.^11^ We sampled the mean number of offspring per mother based on historical Dutch fertility rates, accounting for overdispersion.

**Figure 2:**
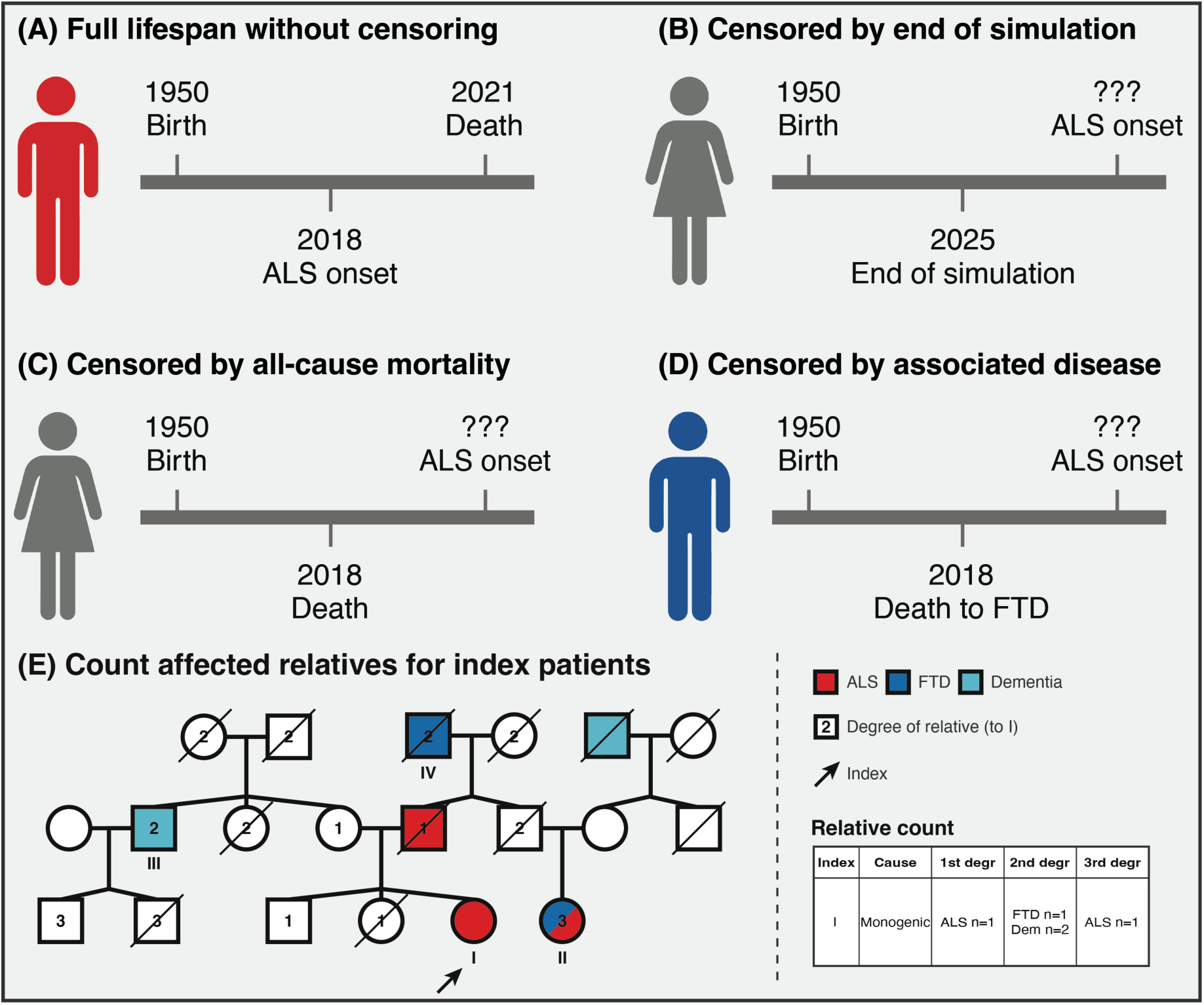
Censoring of phenotypes and counting affected relatives. **(A)** Individual with full lifespan, with ALS onset (red) before death. **(B)** Individual whose potential affected disease status was censored by the end of the simulation. **(C)** Individual whose potential affected disease status was censored by the competing all-cause mortality, due to sampled life expectancy. **(D)** Individual whose potential ALS-affected disease status was censored by the competing mortality from ALS-associated disease (FTD, blue). **(E)** Counting of affected relatives in index patient. Index patient (individual I) is marked by the black arrow. The degree of relatives to individual I is illustrated by the number in each individual. Comorbid ALS-FTD (individual II) is only counted as ALS once. Note that for ‘any dementia’-affected relative, both FTD-(individual IV) and other dementia-affected (individual III) relatives are considered.

#### Monogenic disease model

Supplementary Table 1 summarises the parameters used to model monogenic and polygenic disease with relevant estimates from literature.^8,9,12,13,18^ In the monogenic model, we modelled three classes of pathogenic alleles: ‘*ALS-FTD-moderate* penetrance’ alleles (e.g. *C9orf72*-like), rare, but more penetrant ‘*ALS-FTD-high* penetrance’ alleles (e.g. *FUS/SOD1*-like), and rare, highly penetrant ‘*FTD-specific’* alleles (e.g. *GRN/MAPT*-like). Individuals in the founder generation were assigned genotypes based on population disease allele frequencies or aggregated, gene-specific, pathogenic disease allele frequencies derived from gnomAD v4.1.0 (GRCh38).^8^ Monogenic ALS and FTD were considered autosomal dominant with allele-specific penetrance estimates to determine affected status.

#### Polygenic disease model

In the polygenic model, we used the liability threshold model, in which we simulated a disease liability for all individuals.^30^ The genetic (*G*) and non-genetic (*E*) components for ALS, FTD, and dementia were modelled using heritability (*h*^2^) and genetic correlation (*r_g_*) estimates.^14,15,19,20,22–25^ The liability threshold (*lt*) was defined by the population lifetime risk (*K*). Overall polygenic lifetime risk parameters were corrected for the proportion of disease attributable to polygenic disease (85% in ALS, 75% in FTD).^1,31^

#### Defining disease status

We assigned the final monogenic or polygenic disease status per individual only if disease occurred before censoring in the simulation, accounting for all-cause mortality (i.e. general life expectancy) and competing mortality from other diagnoses (Fig. 2A-D).^11^ We used published data on disease-specific age at onset to sample at what age an individual would have their disease onset (Supplementary File 1A).^13,16,17,21,22^ Competing risk was modelled using disease-specific survival estimates (Supplementary File 1A).^26–29^ If the disease were to occur after death, an individual is considered ‘unaffected’ in the family history for this disease.

#### Simulation benchmarking

Main simulation outputs were validated against input parameters and population-level epidemiological benchmarks, including fertility rate, life expectancy, trait-specific lifetime risks, proportions of monogenic and polygenic disease, allele-specific penetrance, heritability, and familial recurrence rates.

### Statistical analysis

#### Evaluation of fALS criteria

We simulated 1,000,000 three-generation pedigrees. The predictive performance of the fALS criteria was assessed in ALS index patients with disease onset between 2010 and 2025, reflecting current ALS databases. For each index patient, the number of affected relatives within the third degree was counted under three definitions: relatives with (1) only ALS; (2) ALS and/or FTD; or (3) ALS and/or any dementia (Fig. 2E). To reflect a family history, we only scored affected relatives at the time of disease onset.

We first examined the accuracy of fALS criteria proposed in earlier studies,^3,4^ by calculating the positive predictive value (PPV), false positive rate (1-PPV), and sensitivity for detecting monogenic disease. We then assessed the added predictive value of considering relatives with FTD or any form of dementia. We also examined the predictive effect of considering more distant relatives up to the third degree.

To improve upon traditional fALS criteria, we evaluated a series of logistic regression models. Using 1,000,000 family histories, the probability of monogenic ALS was modelled as a function of the number of affected and unaffected relatives (Supplementary File 1B). We cross-validated the three models scoring either ALS, ALS-FTD, or ALS and all-cause dementia using the *caret* R package with tenfold cross-validation. We then tested the model’s predictive performance in an independent set of 1,000,000 simulated pedigrees.

#### Sensitivity analyses

Sensitivity analyses for the following parameters were included: fertility, life expectancy, age at monogenic and polygenic disease onset, disease allele frequencies, allele-specific penetrance, lifetime risk, heritability, and genetic correlation, as well as year of simulation censoring effects (Supplementary Table 1, Supplementary Table 2). For our sensitivity analyses, we performed tenfold simulations of at minimum 100,000 pedigrees. Sensitivity and PPV were reported as a mean ± 1 standard deviation (SD) over 10 repetitions.

#### Shiny application

We built a Shiny application based on three-generation pedigree simulations, available at https://paulbeele-apecs-monogenic-probability-calculator.share.connect.posit.cloud/. In this app, we provide the PPV and 95% confidence interval of having up to five relatives with ALS/FTD, up to third degree relatives, based on 50,000,000 simulations with the ‘main simulation’ parameters. In addition, the ‘variable parameters’ tab allows for adjustment of the monogenic ‘*ALS-moderate*’ (range 10% - 50%) and ‘*ALS-high*’ (range 30% - 70%) allele ALS penetrances, as well as the polygenic ALS heritability (range 20% - 60%). The variable parameters tab provides PPV and 95% confidence intervals over 10,000,000 simulations of each parameter combination.

#### Phenocopy rate

For the main simulations, we calculated the proportion of monogenic index cases with relatives with discordant causes of ALS, i.e. “phenocopies”. We defined phenocopies as relatives with either polygenic ALS, or relatives with monogenic ALS carrying a pathogenic disease allele they inherited from a different ancestor than the index patient.

#### Cryptic distant relatedness

We ran 50,000 four-generation simulations where the founder always carried the *ALS-FTD-moderate* penetrance disease allele. We then quantified apparently sporadic monogenic cases, defined as having no ALS or FTD among first- and second-degree relatives, and cousins (third-degree relatives; Supplementary Fig. 1, grey area). We then assessed the added value of extending the family history to include further third-, fourth-, or fifth-degree relatives (Supplementary Fig. 1, white area).

## Results

### Simulation benchmarking

Simulating 1,000,000 three-generation pedigrees resulted in 5,431 index patients with ALS. Of these, 893 (16.4%) had monogenic ALS and 4,538 (83.6%) had polygenic ALS. In total, 181,835 relatives were scored for the family history. Of these relatives, 1,947 had ALS, 527 had FTD, and 3,226 had any form of dementia. The full lifespan analyses indicate that the simulations are unbiased and well-calibrated as the estimates derived from simulated pedigrees reflect input parameters and real-world demographics (Table 1, Supplementary Fig. 2).

**Table 1:**
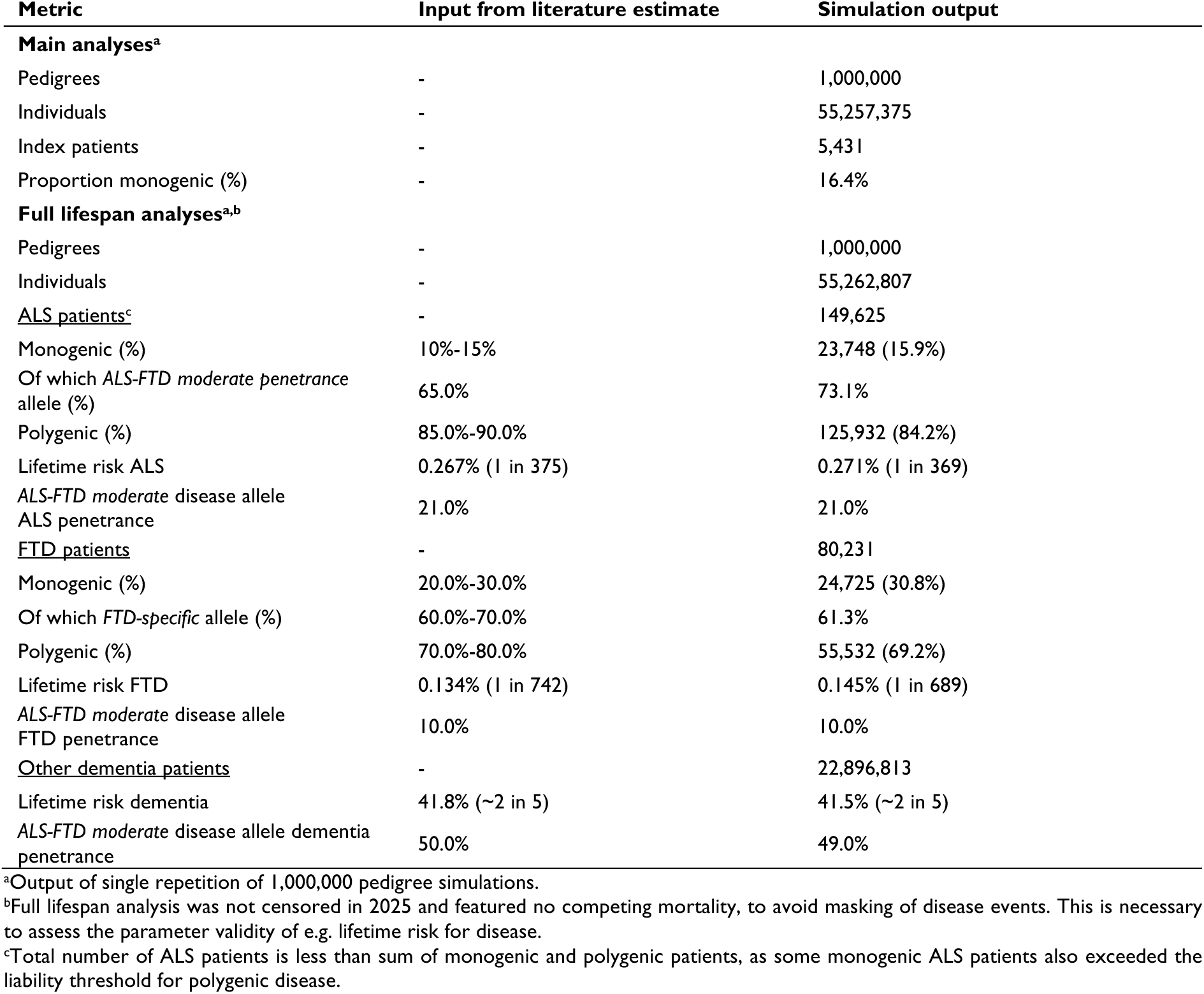
Benchmarking of input parameters and literature estimates to simulation output.

Supplementary File 1A illustrates that the demographic estimates based on simulation output, age at onset, and survival after onset are unbiased estimates of the input parameters. Supplementary Fig. 2 illustrates that monogenic and polygenic simulation output matches input parameters, regarding Mendelian autosomal disease inheritance (Supplementary Fig. 2A), polygenic recurrence risk (Supplementary Fig. 2B), polygenic heritability (Supplementary Fig. 2C), and lifetime risk (Supplementary Fig. 2D).

### Evaluation of fALS criteria

In total, 23.6% of the patients with ALS had at least one relative with ALS and/or FTD up to the third degree (Fig. 3). While patients with polygenic ALS had fewer affected relatives, still 12.5% of polygenic cases had ≥1 relative with ALS or FTD within the third degree. Conversely, 24.0% and 8.5% of monogenic ALS patients with the *moderate* or *high* penetrance disease allele, respectively, had no ALS- or FTD-affected relatives within the third degree.

**Figure 3:**
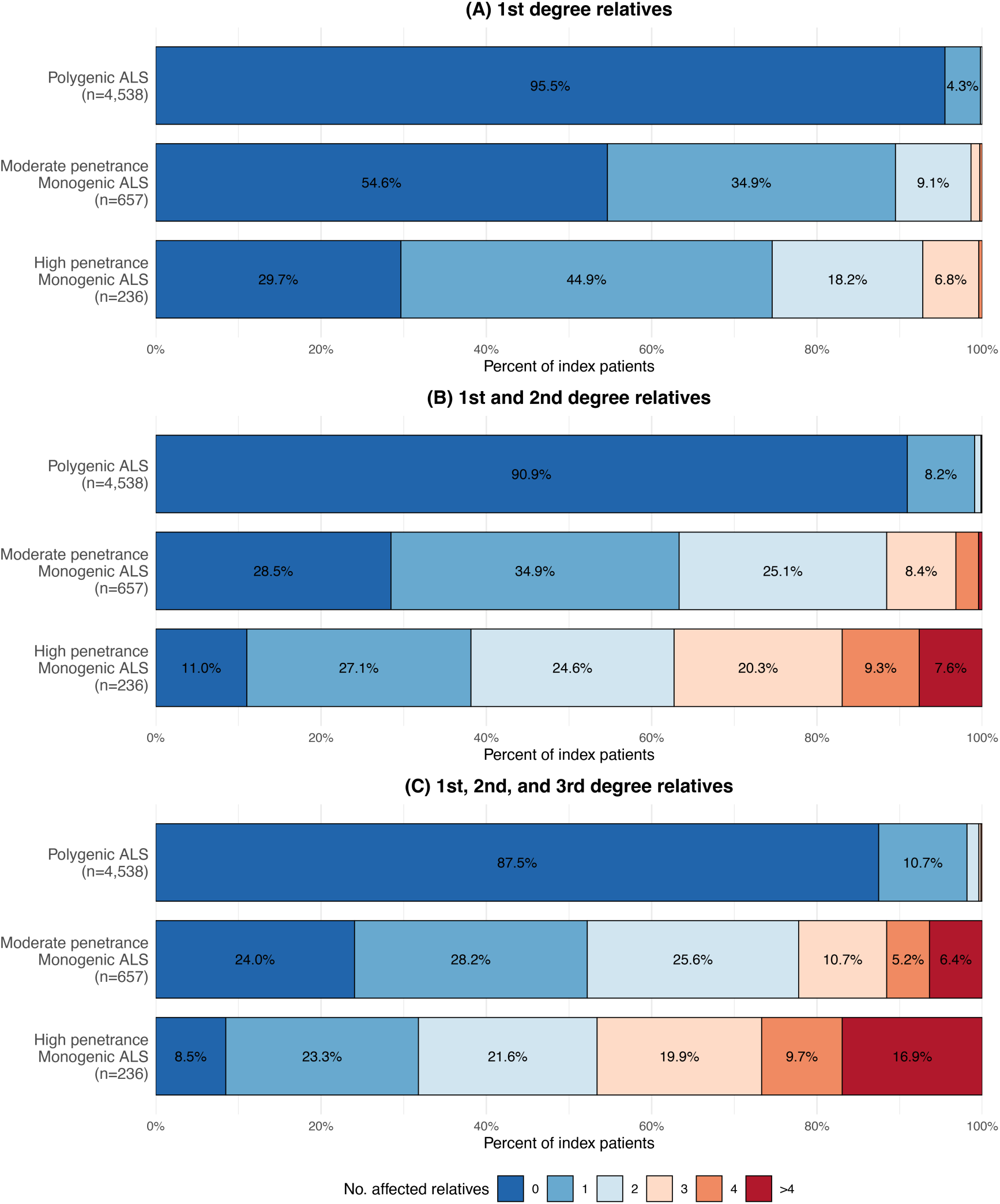
Distribution of number of ALS or FTD-affected relatives for index patients. Proportion of index patients with polygenic ALS, moderate penetrance monogenic ALS, or high penetrance monogenic ALS who have *n* ALS- or FTD-affected relatives up to the third degree.

Fig. 4 illustrates the difference between clinicians’ assessment of familial ALS and the positive predictive value for having monogenic ALS, previously enquired by Byrne *et al*.^3^ Clinicians tended to overestimate the proportion of monogenic disease if the index patient had a first-degree affected relative (Fig. 4A-B), while being more accurate estimating the proportion of monogenic disease if only second-degree relatives were affected (Fig. 4C-D).

**Figure 4:**
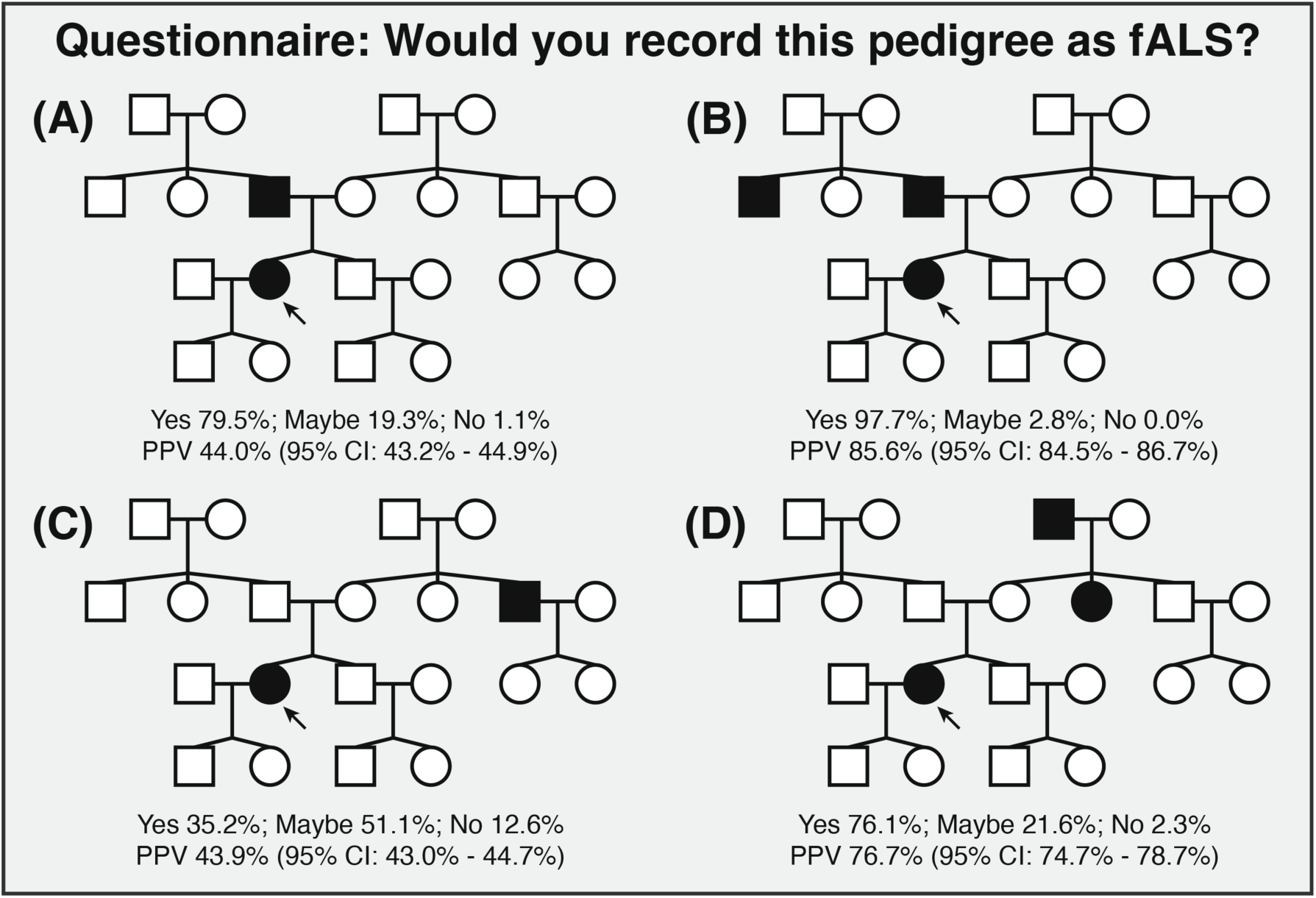
Benchmarking of clinicians’ estimates of “familial ALS”. Pedigrees and questionnaire results were derived from Byrne *et al.*^3^ on the lack of fALS definition consensus. Index patient marked by arrow. PPV: positive predictive value for having underlying monogenic disease as cause of ALS. 95% CI: 95% confidence intervals.

Table 2 demonstrates the predictive accuracy of various fALS criteria cutoffs. The proposed criterion for ‘probable fALS’ (having one first/second degree relative with ALS) had a PPV of 48.6%, misclassifying 51.4% of patients as monogenic. The proposed criterion for ‘definite fALS’ (having at least two first/second degree relative with ALS) increased PPV to 90.0%, at the cost of decreasing sensitivity to 31.1%, missing 68.9% of monogenic patients. The sensitivity increases to 43.3% by taking ALS and FTD into account (≥2 relatives with ALS or FTD). Considering ALS- or FTD-affected relatives up to the third-degree further improved sensitivity, while slightly decreasing the PPV. Dependent on the exact criteria, 9.8% - 15.0% of ‘familial ALS’ patients would falsely be classified as monogenic, while missing 46.8% - 56.7% of monogenic ALS index patients, that did not reach the fALS criterion cutoff. Considering any form of dementia markedly increased the false positive rate, when trying to predict monogenic disease. The Shiny app available at https://paulbeele-apecs-monogenic-probability-calculator.share.connect.posit.cloud/ provides further PPV estimates over a range of specific ALS/FTD affected relative combinations.

**Table 2:** Predictive accuracy of fALS criteria.

| fALS criteria | Sensitivity<sup>a</sup> | Positive predictive value<sup>a</sup> | False positive rate<sup>a</sup><br>(1 - PPV) |
| --- | --- | --- | --- |
| **Relatives within second degree** |  |  |  |
| 1 relative with ALS<sup>b</sup> | - | 48.6% (44.8%–52.4%) | 51.4% (47.6%–55.2%) |
| ≥1 relative with ALS | 66.2% (63.0%–69.2%) | 62.0% (58.9%–65.0%) | 38.0% (35.0%–41.1%) |
| ≥2 relatives with ALS | 31.1% (28.2%–34.2%) | 90.0% (86.2%–92.8%) | 10.0% (7.2%–13.8%) |
| ≥2 relatives with ALS or FTD | 43.3% (40.1%–46.6%) | 90.2% (87.1%–92.6%) | 9.8% (7.4%–12.9%) |
| ≥2 relatives with ALS or dementia | 63.5% (60.3%–66.6%) | 64.1% (60.9%–67.2%) | 35.9% (32.8%–39.1%) |
| **Relatives within third degree<sup>c</sup>** |  |  |  |
| ≥1 relative with ALS | 71.7% (68.6%–74.5%) | 56.1% (53.2%–59.0%) | 43.9% (41.0%–46.8%) |
| ≥2 relatives with ALS | 40.4% (37.3%–43.7%) | 85.1% (81.5%–88.2%) | 14.9% (11.8%–18.5%) |
| ≥2 relatives with ALS or FTD | 53.2% (49.9%–56.4%) | 85.0% (81.8%–87.7%) | 15.0% (12.3%–18.2%) |
| ≥2 relatives with ALS or dementia | 70.1% (67.0%–73.0%) | 52.3% (49.5%–55.1%) | 47.7% (44.9%–50.5%) |

Predictive accuracy of various fALS definitions predicting monogenic inheritance based on number of affected relatives of an ALS-affected index patient in three-generation pedigrees.
<sup>a</sup>Values shown as: predictive measure percentage (95% Confidence Intervals).
<sup>b</sup>Sensitivity cannot be calculated for fALS criteria matching an exact number of relatives as pedigrees with both more and less than the number of affected relatives do not match the criterion.
<sup>c</sup>Three-generation pedigrees allow for cousins (children of uncles/aunts) as third degree relatives. See Supplementary Figure 1.

The logistic regression models used for predicting monogenic diseases are detailed in the Supplementary File 1B. Inclusion of more distant relatives, as well as considering FTD or dementia-affected relatives improved the models’ ability to distinguish monogenic from polygenic disease (Supplementary Fig. 3A-B). Notably, the discriminatory performance increased when the model included both ALS and dementia-affected relatives, compared to the model restricted to ALS and definite FTD (Supplementary Fig. 3B), attributable to the higher sensitivity achieved by encompassing the more prevalent dementia cases. The prediction models incorporating both the affected and unaffected relatives, achieved the highest discriminatory performance (Supplementary Fig. 3C).

After cross-validating the models utilising both affected and unaffected relatives, we applied the trained prediction model to index patients of the main simulation. Discriminatory performance was comparable to the training set (Supplementary Fig. 4). Next, we set the model’s threshold for predicting monogenic disease to match the sensitivity of the classical fALS criteria for comparison. The prediction models showed a minor improvement in PPV upon the dichotomous fALS criteria when predicting underlying monogenic disease (Supplementary Table 3).

fALS criteria considering ≥3 affected relatives with ALS/FTD reached a PPV ≥93.1%, serving as reliable predictors for definite monogenic disease. Dependent on the exact criteria, at most 5.1% of pedigrees had this many affected relatives up to the third degree (Supplementary Table 4).

### Sensitivity analyses

We repeated analyses with varying values for simulation parameters (Supplementary Table 2, Supplementary Fig. 5). We set the fALS criterion for sensitivity analyses to predict monogenic inheritance at having ≥2 affected relatives with ALS or FTD within the second degree, matching the proposed criterion for ‘definite fALS’ incorporating FTD.^4^

Sensitivity of the fALS criteria decreased with decreasing fertility rates, as small pedigrees impeded familial clustering of disease (Supplementary Fig. 5A). When the mean life expectancy exceeded 70 years, the accuracy of the fALS criterion remained relatively stable (Supplementary Fig. 5B). Historically, this happened after the 1940s (Supplementary File 1A). Since monogenic ALS had an earlier onset than polygenic ALS (Supplementary File 1A), the fALS criterion sensitivity decreased when monogenic ALS had a later onset, with less familial clustering at a later age due to competing mortality (Supplementary Fig. 5C). Similarly, the PPV improved when polygenic disease occurred later (Supplementary Fig. 5D). If the ratio of simulated monogenic to polygenic ALS index patients shifted towards more monogenic index patients, through increased disease allele frequency, increased disease allele penetrance, or decreased polygenic lifetime risk, predictive accuracy improved (Supplementary Fig. 5E-I). Of note, an *ALS-FTD-moderate* disease allele penetrance of 30%, which is already a relatively high estimate compared to *C9orf72* penetrance estimations in literature, had moderate predictive performance. While it improved the PPV to 93.3%, sensitivity remained only 49.8%, missing half of all monogenic cases. Higher ALS heritability resulted in more familial clustering of polygenic ALS and FTD disease, decreasing the fALS criterion PPV (Supplementary Fig. 5J). Finally, the sensitivity of the fALS criteria is projected to decrease in the future as pedigree sizes decrease, reflecting the combined effect of increasing life expectancies and decreasing fertility rates (Supplementary Fig. 5L).

### Phenocopy rate

Among 893 monogenic index cases, 47 patients (5.3%) had at least one relative with polygenic ALS or monogenic ALS with the disease allele originating from a different ancestor, instead of concordant *ALS-FTD-moderate* or *ALS-FTD-high* penetrance disease allele carriership. Phenocopies occurred more frequently in more distant relatives of monogenic index patients, ranging from 0.6% (three out of 496) in the first degree to 5.2% (28 out of 544), and 7.9% (26 out of 330) of ALS-affected relatives in the second and third degree, respectively.

### Cryptic distant relatedness

We expanded the three-generation pedigree simulations to 50,000 four-generation pedigree simulations, with each core pedigree founder carrying the *ALS-FTD-moderate* penetrance disease allele. This allowed for counting more distant relatives (Supplementary Fig. 1). Among monogenic index patients (*n* = 11,730), 2,252 patients (19.2%) had no ALS- or FTD-affected relatives up to cousins (third-degree relatives, Supplementary Fig. 1, grey area). Of these apparently sporadic pedigrees, 1,871 index patients (83.1%) had at least one distant ALS- or FTD-affected relative in the third (*n* = 1,262; 56.0%), fourth (*n* = 1,335; 59.3%) or fifth (*n* = 1,077; 47.8%) degree.

## Discussion

In this study, we assessed the predictive utility of proposed clinical criteria for familial ALS to distinguish between monogenic and polygenic ALS. From empirical demographic data, we simulated pedigrees under both monogenic and polygenic models of ALS and related disorders. Our findings build on prior studies challenging the traditional familial-sporadic dichotomy derived from clinical history alone. Previously, it was shown that complex diseases are typically sporadic but can manifest familially,^5^ while monogenic ALS may appear sporadic, particularly in small nuclear pedigrees with incomplete penetrance.^6^ By integrating these perspectives into a unified framework across multi-generational pedigrees, we address the ongoing lack of consensus on fALS definitions.^3,4^ We demonstrate that a family history suggestive of fALS does not necessarily indicate monogenic inheritance.

Our results suggest that the fALS criteria systematically overestimate the proportion of pedigrees resulting from a single, monogenic cause. Under the ‘best’ fALS definition (two or more relatives with ALS or FTD within the second or third degree) approximately one in seven to ten fALS pedigrees results from clustered polygenic disease. This false positive rate further increases when including families with only one ALS- or FTD-affected relative, or when misclassifying any form of dementia as ‘definite FTD’. Clinicians need to be aware of this and discuss genetic risk thoughtfully to avoid causing unnecessary alarm, recognising that a substantial proportion of pedigrees fulfilling fALS criteria may display polygenic inheritance. From a research perspective, these polygenic fALS pedigrees cannot be ‘resolved’ by the identification of a single pathogenic variant as by definition there is none,^32^ as clustering resulted from shared polygenic liability.

We found that when an index patient has three or more affected relatives with ALS or FTD, a monogenic aetiology can be reliably suspected (PPV ≥93.1%). In our simulations, however, at most 5.1% of index patients had this many affected relatives. While these cutoffs might serve as a reliable proxy for inferring underlying monogenic disease, in clinical practice, these fALS criteria can only be applied to a small subset of all ALS patients.

Importantly, our simulations show that clinicians cannot assume that sporadic ALS patients lack a monogenic cause. The identification of an underlying variant might increasingly inform prognosis and, in the future, therapeutic eligibility. A sporadic presentation does not preclude a monogenic origin, particularly as decreasing family sizes will increasingly obscure inheritance patterns, causing monogenic disease to present as isolated cases. As such, comprehensive genetic testing needs to be part of routine clinical evaluation to avoid missing potentially treatable cases.^2^ Our simulations indicate that 83.1% of apparently sporadic, monogenic index patients had cryptic, distant relatives with ALS or FTD up to the fifth degree. Future research can thus also benefit from exploring methods to uncover distant relatedness and shared ancestral variants, such as through regional family registries or identity-by-descent detection tools, to resolve the monogenic subset of patients currently classified as sporadic.^33^

Finally, our study demonstrates that ALS can manifest under both monogenic and polygenic models within the same pedigree, resulting in so-called phenocopies. A previous study in *C9orf72*-positive Irish kindreds identified ALS-affected, non-*C9orf72* patients, reporting an elevated lifetime risk for non-carriers in *C9orf72* pedigrees.^34^ Here we extend this observation, finding ‘phenocopy’ relatives in 5.3% of monogenic index patients, reflecting the high population prevalence of polygenic disease and probability of co-occurrence. This underscores the need for comprehensive genetic testing across all affected pedigree members, rather than assuming a uniform cause of ALS within a pedigree, especially in the era of gene-targeted treatment for all patients in a pedigree.

Our study has several limitations. Our simulation adopted a simplified dichotomy between monogenic and polygenic inheritance, whereas complex disease in ALS may also involve oligogenic contributions from a small number of medium-effect variants, with odds ratios deviating from polygenic normality.^1,35^ These oligogenic variants drive familial clustering beyond standard polygenic risk. However, reliable parameter estimates for simulating oligogenic variants, such as the number of variants, their allele frequencies, and their effect sizes are lacking. This hampers reliable modelling. Speculatively, substituting a fraction of our polygenic index cases (comprising 83.6% of index cases) with more familially clustered, oligogenic pedigrees would likely further impair the predictive accuracy of the fALS criteria for strictly monogenic disease. Future work might expand upon our current framework by considering a more complex architecture.

A second limitation concerns the reliance on simulations to evaluate fALS criteria performance. Although simulation-based approaches are inherently imperfect, unbiased real-world data on family histories and corresponding genetic outcomes remain unavailable. Clinicians’ thoroughness in ascertaining family history correlates with prior suspicion of monogenic disease, while patient-reported histories become unreliable for distant relatives. Moreover, no gold-standard genetic testing paradigm exists, and the pathogenicity of variants of uncertain significance is rarely immediately clear in practice.^10^ By contrast, reliable parameters for ALS and related disorders under both monogenic and polygenic models are robustly established, strengthening our approach.^7–9,11–29^

Despite these limitations, our simulation outputs closely mirrored literature estimates on demographic and genetic ALS data. Although optimal input parameters may vary with emerging disease insights, sensitivity analyses robustly confirmed the imperfection in fALS criteria to detect monogenic disease over a range of plausible parameter values. Ultimately, the simulations likely still overestimated true predictive accuracy, owing to perfect ascertainment of family history up to third-degree relatives in simulations, unlike in an imperfect, patient-reported real-world context.

In conclusion, a family history provides limited accuracy for inferring genetic architecture in ALS. Relying on traditional fALS criteria risks both missing cases for genetic testing and treatments, and unnecessarily alarming families with polygenic ALS. Therefore, we propose to estimate the probability of monogenic disease as a continuum, based on a family history. Given the limited value of a family history and risk of phenocopies, routine genetic testing regardless of family history and systematic approaches for detecting distant relatedness will help to find known and novel genetic causes in ALS.

## Data availability

All simulations and analyses were conducted using R version 4.4.2. The data and code used for this study will be available from Github upon publication: https://github.com/wvanrheenen/APECS.

## Acknowledgements

Authors have no further acknowledgements to declare.

