## Supplementary Figures for "The value of a family history to distinguish monogenic from polygenic amyotrophic lateral sclerosis"

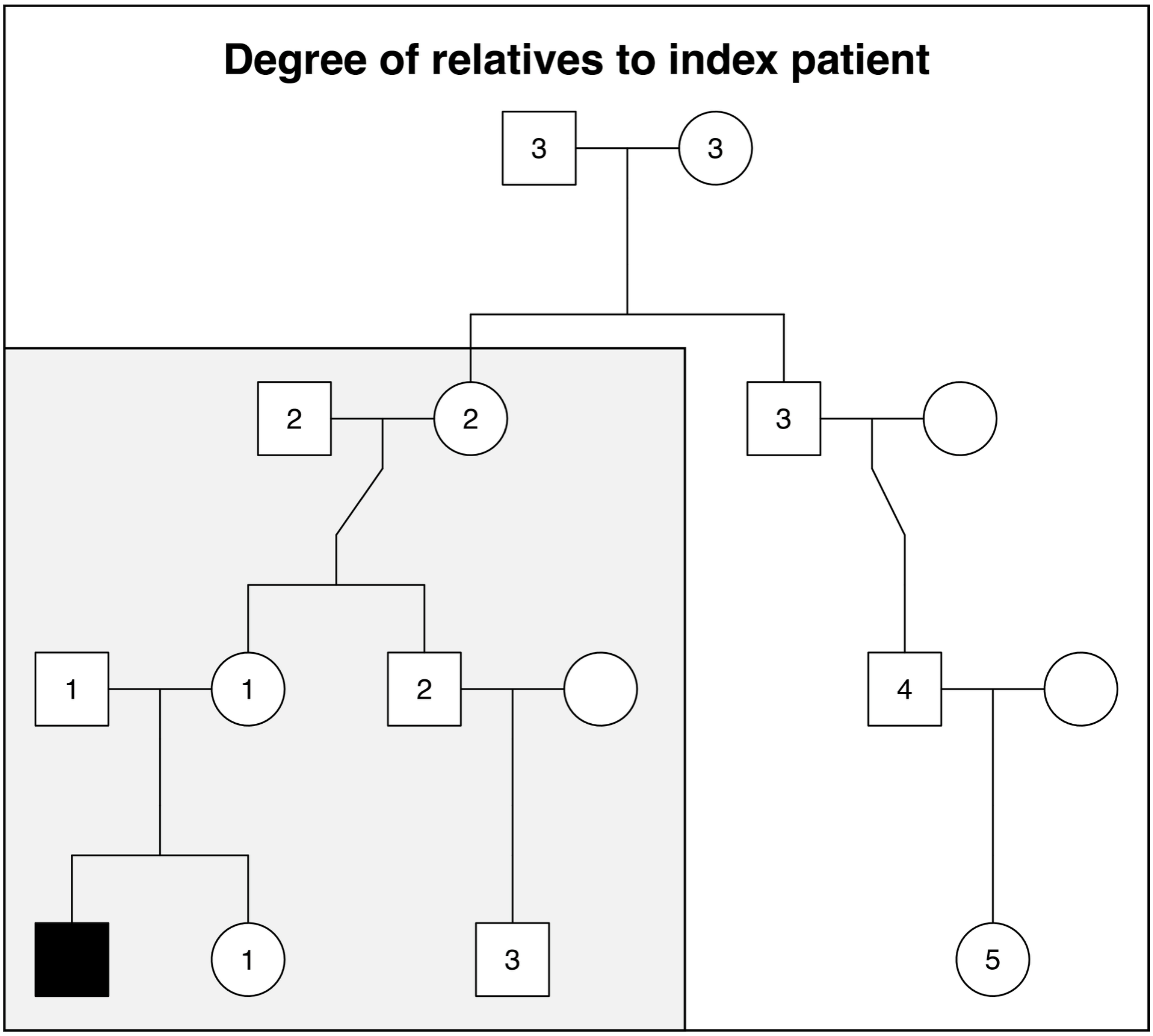


**Supplementary Figure 1: Degree of distant relatives to index patients.** Index patient is marked in black. In grey, the degree of relatives in three-generation pedigrees. Note how cousins are marked as ‘close’ third-degree relatives. In white, the degree of distant relatives in four-generation pedigrees.

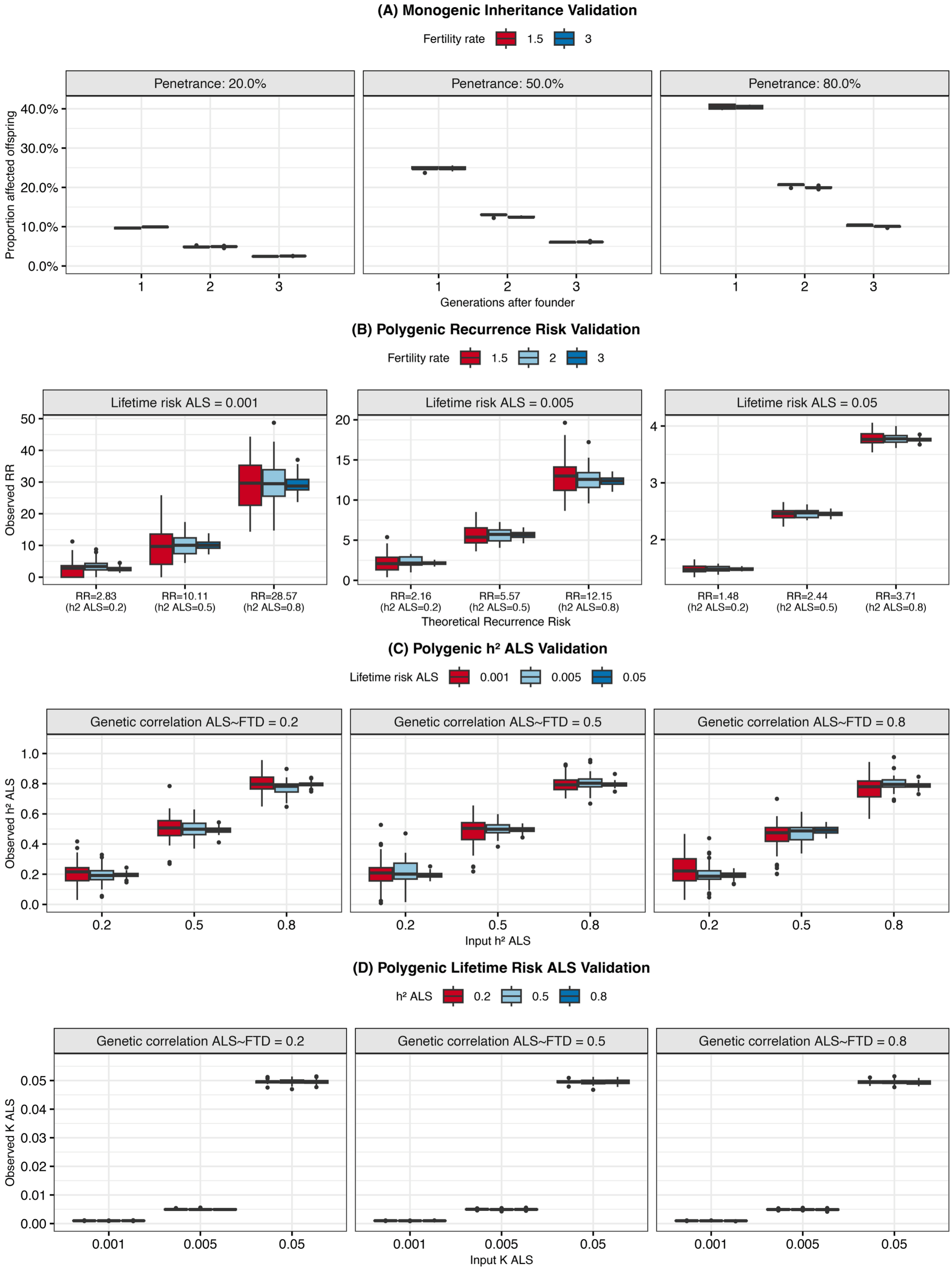


**Supplementary Figure 2: Benchmarking of phenotypic disease parameters. (A) Monogenic autosomal dominant inheritance validation.** Simulated proportion of offspring of an affected founder carrying a monogenic disease allele for various penetrances. Each generation the proportion of affected individuals halves, compared to the prior generation.

**(B) Validation of polygenic recurrence risk.** Simulated vs. theoretical recurrence risk (RR) of first-degree relatives of polygenic ALS patients being affected, according to Falconer's theorem. **(C) Polygenic heritability validation.** Simulated vs. input heritability ($h^2$) of polygenic ALS for various ALS lifetime risks ($K$) and genetic correlation values ($r_g$). **(D) Polygenic lifetime risk validation.** Simulated vs. input lifetime risk ($K$) of polygenic ALS for various ALS heritabilities ($h^2$) and genetic correlation values ($r_g$).

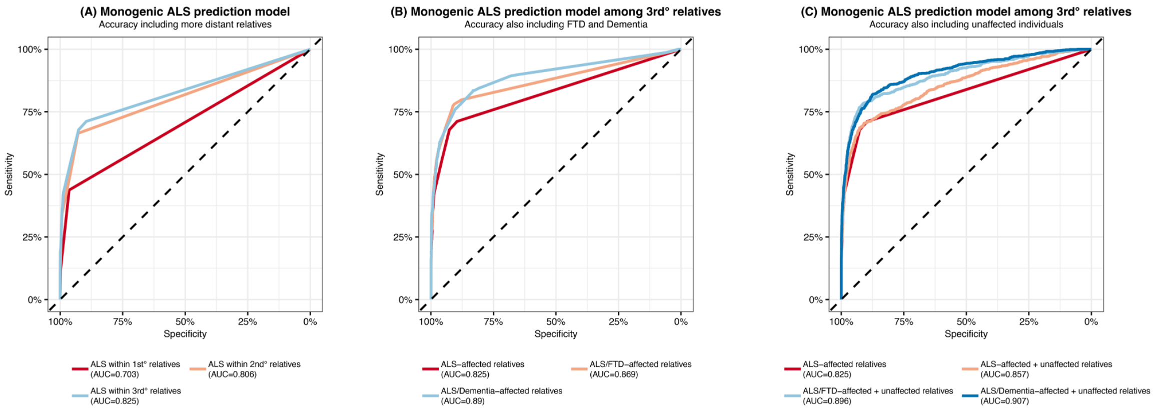


**Supplementary Figure 3: Predictive accuracy of logistic regression models in training set**. (**A)** ROC-curves of models predicting monogenic inheritance based on having ALS-affected relatives within the first, second, or third degree. (**B**) ROC-curves of models predicting monogenic inheritance based on having relatives within the third degree, with ALS, FTD, or any dementia. (**C**) ROC-curves of models predicting monogenic inheritance based on having relatives within the third degree, with ALS, FTD, any dementia, and the number of unaffected relatives.

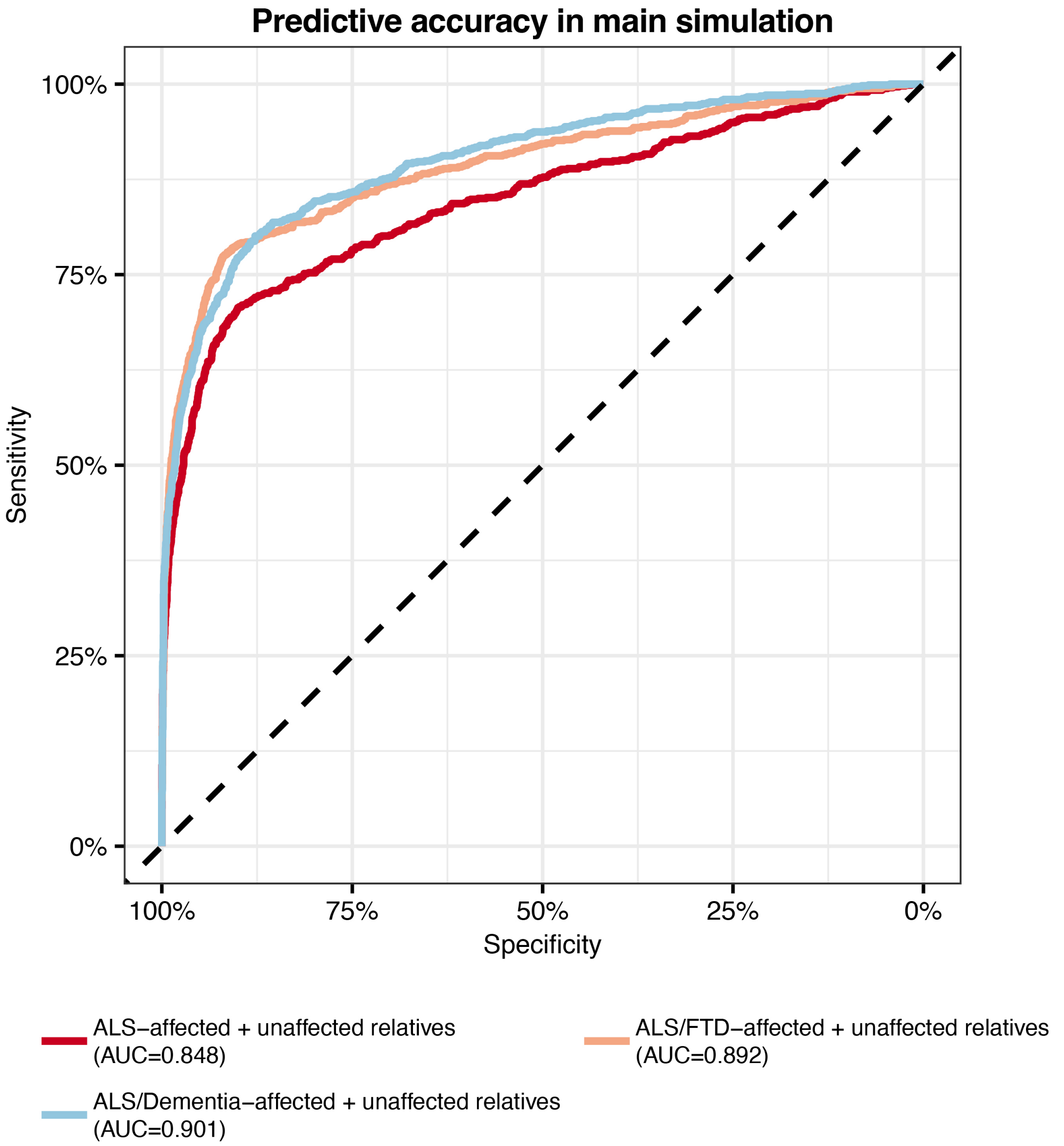


**Supplementary Figure 4: Predictive accuracy of logistic regression models in main simulation test set**. ROC-curves of trained prediction models predicting monogenic inheritance in index patients in the main simulation ($N$ = 5,431) based on having relatives with ALS, FTD, any dementia, and the number of unaffected relatives within the third degree. The prediction model was trained in 1,000,000 simulated pedigrees and then tested on the 5,431 index patients from the 1,000,000-pedigree main analysis.

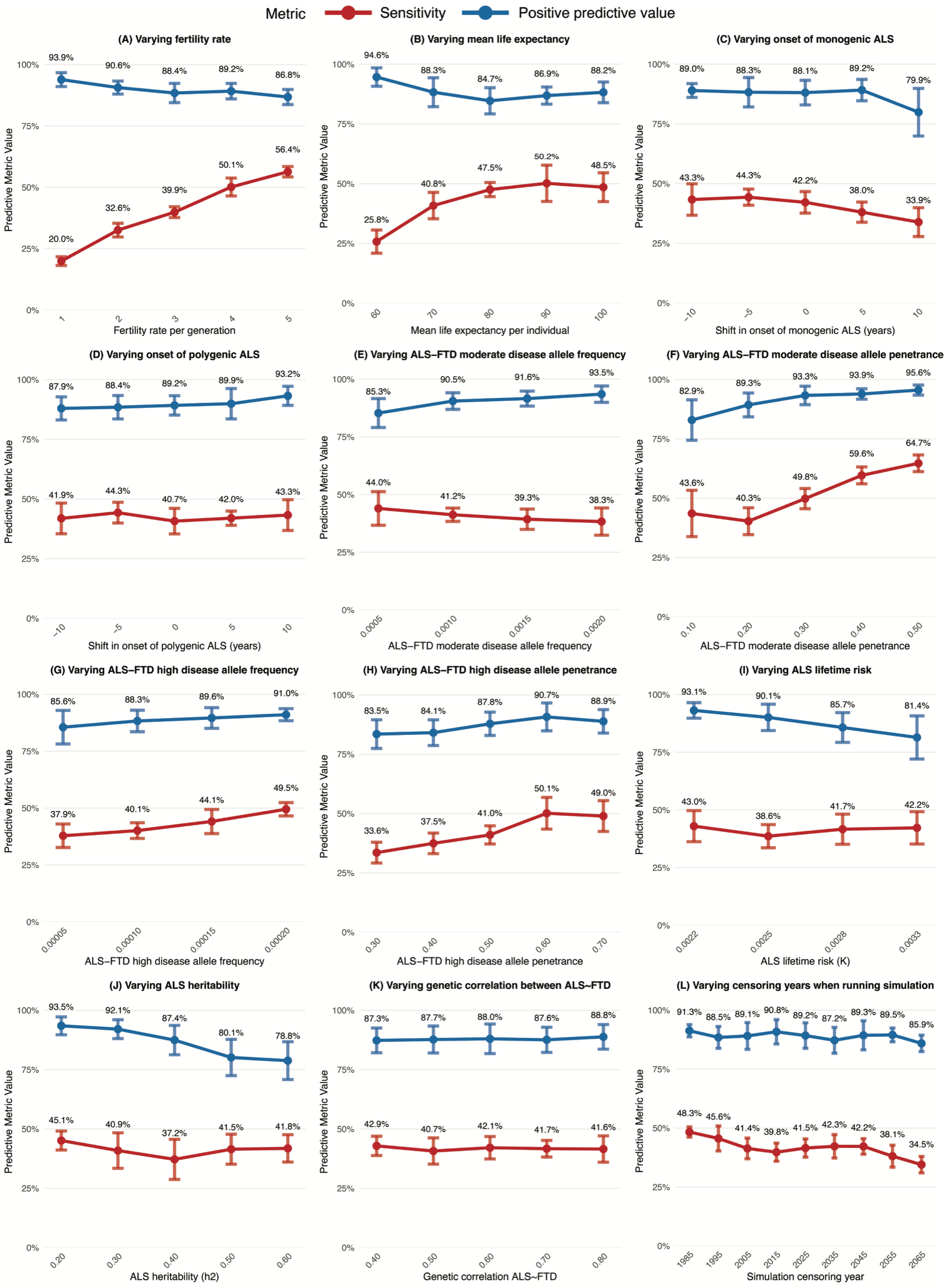


**Supplementary Figure 5: Sensitivity analyses of fALS criteria with varying input parameter values**. Sensitivity and positive predictive value of an index patient having ≥2 relatives with ALS or FTD within the second degree as a predictor for underlying monogenic disease. **(A)** Predictive accuracy for varying fertility rates (1 – 5 children), **(B)** for varying life expectancies (60 – 100 years), **(C)** for shifting the age at monogenic ALS onset (±10 years), **(D)** for shifting the age at polygenic ALS onset (±10 years), **(E)** for varying *ALS-FTD moderate* penetrance disease allele frequency (0.05% – 0.20%), **(F)** for varying *ALS-FTD moderate* penetrance disease allele penetrance (10% – 50%), **(G)** for varying *ALS-FTD high* penetrance disease allele frequency (0.005% – 0.020%), **(H)** for varying *ALS-FTD high* penetrance disease allele penetrance (30% – 70%), **(I)** for varying ALS lifetime risks (0.22% – 0.33%; 1 in 450 to 1 in 300 people), **(J)** for varying ALS heritabilities (20% – 60%), **(K)** for varying ALS~FTD genetic correlations (40% – 80%), and **(L)** for varying years of simulation censoring (1985 – 2065).
