## Supplementary Methods for "The value of a family history to distinguish monogenic from polygenic amyotrophic lateral sclerosis"

### Supplementary File 1A: APECS pedigree simulations

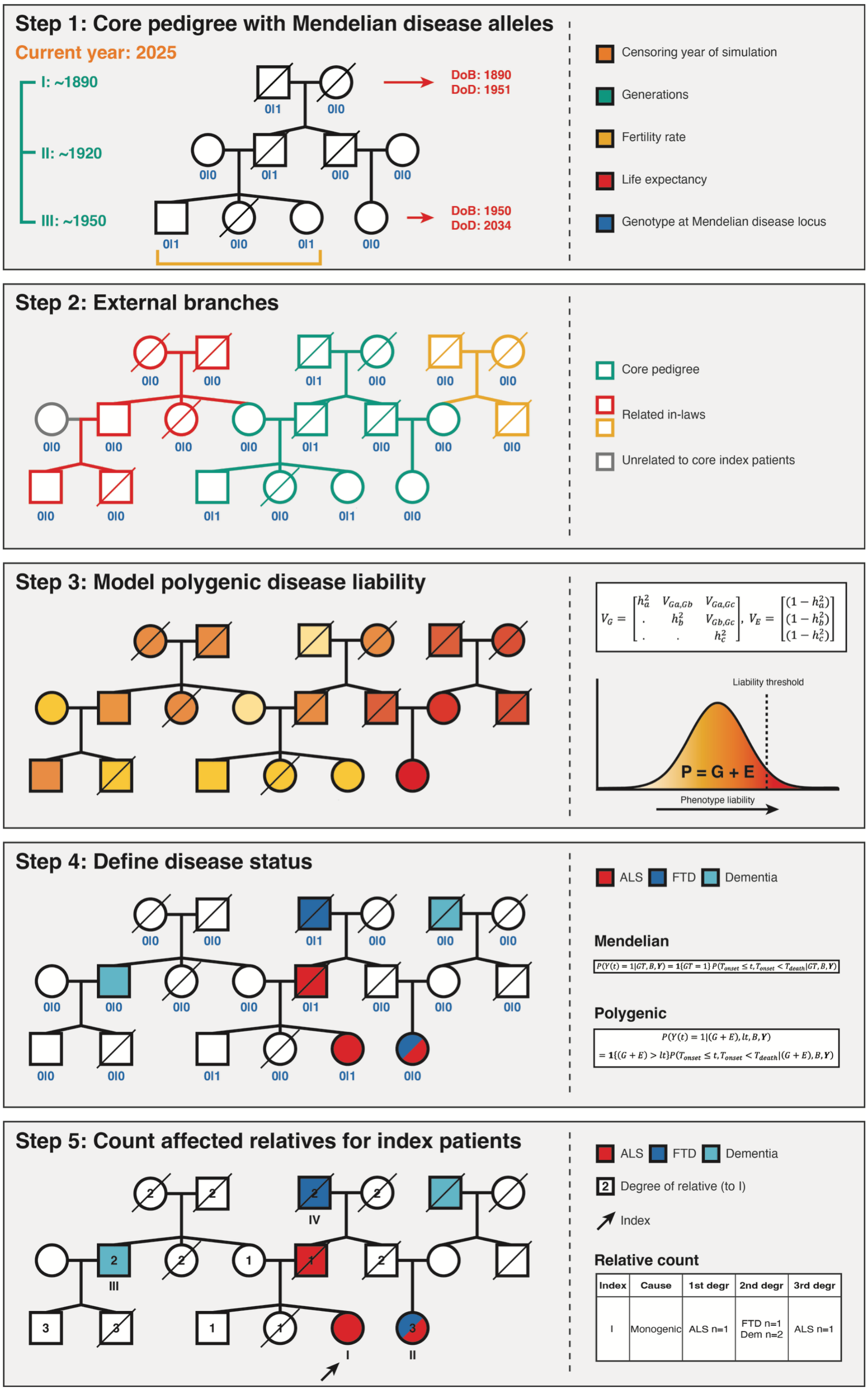

### Step 1: Simulate the core pedigree

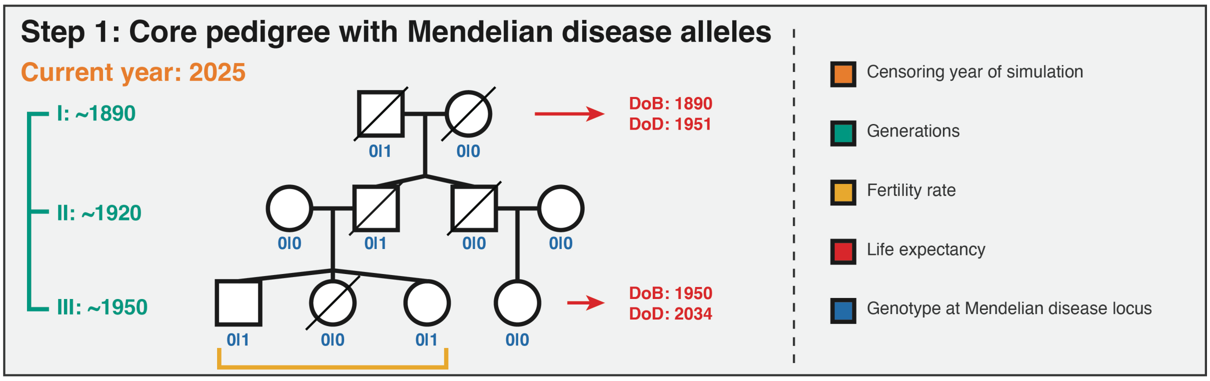


From the user defined index generation year of birth, we calculate *k* generations back where a generation is sampled from a truncated normal distribution with finite intervals between 25 and 35 years. This defines the birthyear of the ancestor pair.

From this date, we simulate an ancestor pair with the life expectancy sampled from a left-skewed normal distribution such that the mean is equal to the historical mean life expectancy for their birthyear (Figure 1).[1] α and κ are set as to mirror a limit to maximum biological age, with a maximum life expectancy of 100 years:

$$X \sim skewNormal(\xi, \omega, \alpha) \qquad (1)$$

$$\xi = historical_{mean\ life\ expectancy} - \omega * \sqrt{\frac{2}{\pi}} * \frac{(\alpha)}{\sqrt{1+\alpha^2}}, with\ \alpha = -3 \qquad (2)$$

$$\omega = \kappa * \frac{100}{historical_{mean\ life\ expectancy}}, with\ \kappa = 5.5 \qquad (3)$$

Initial survival status is defined independently of disease status, using the simulation's censoring year and each patient's date of death. Individuals are considered deceased if the current simulation date is later than their date of death.

### Figure 1: Simulation of life expectancy

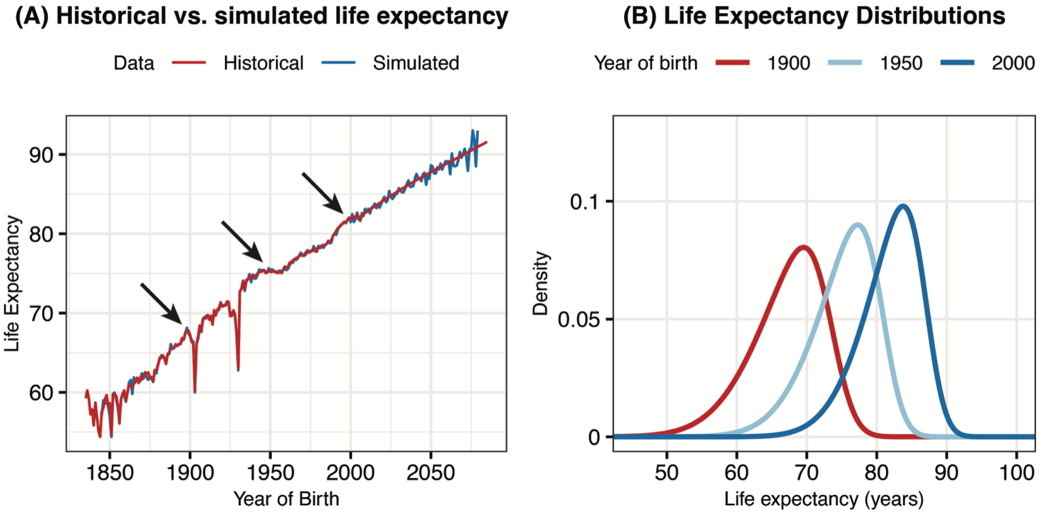


At each Mendelian disease locus, the alleles of the ancestors are modelled as a Bernoulli draw with probability $p$, where $p$ is the user defined disease-allele frequency.

Next, we simulate offspring of the ancestors (second generation) where the number of offspring is sampled from a negative binomial distribution:

$$X \sim NegBin(r, \mu), with\ r = 5, and\ \mu = historical_{mean\ fertility\ rate} \qquad (4)$$

Where the dispersion parameter $r = 5$ and the mean ($\mu$) is defined by the historical fertility rate at the birthyear of the female ancestor plus an intergenerational interval (i.e. offspring birthyear) sampled from a truncated normal distribution with finite intervals between 25 and 35 years (Figure 2).[2]

### Figure 2: Simulation of fertility rates

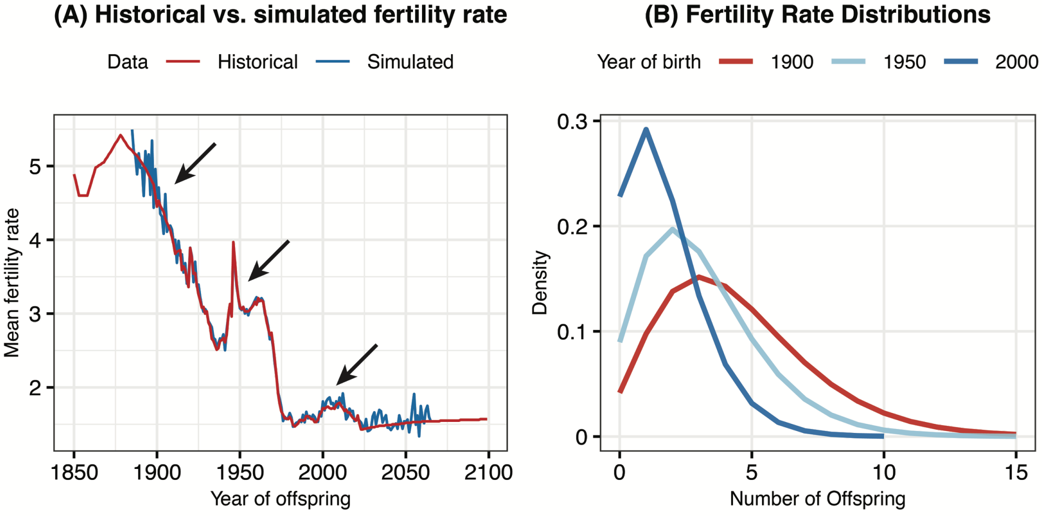

We force at least one offspring for the firstborn child per generation in core pedigree individuals, to guarantee simulation of at least one potential core pedigree index patient in the $k^{th}$ generation. This slightly inflates fertility rates in generations with less offspring.

Offspring receive alleles following the Mendelian law of autosomal segregation. Monogenic ALS and FTD are considered autosomal dominant with reduced penetrance (Supplementary Table 1).[3–7]

Mating partners are simulated in the same way as the ancestors, assuming random mating. Note that in the figure we only display one disease locus, but multiple disease loci for multiple diseases can be defined simultaneously.

### Step 2: Simulate external branches

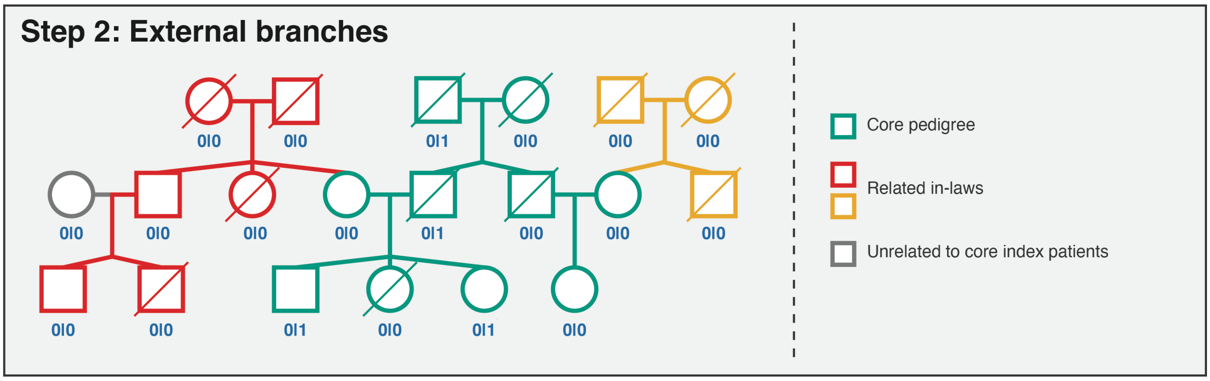


For each partner married into the core pedigree we generate their parents and potential siblings. Next, we simulate offspring for each of the in-laws to generate cousins of the index in case of a three-generation pedigree. For each individual, we model life expectancy and number of offspring. For new founders in in-law branches, we sample alleles of ancestors at each Mendelian disease locus, as described in step 1.

### Step 3: Model polygenic disease liability

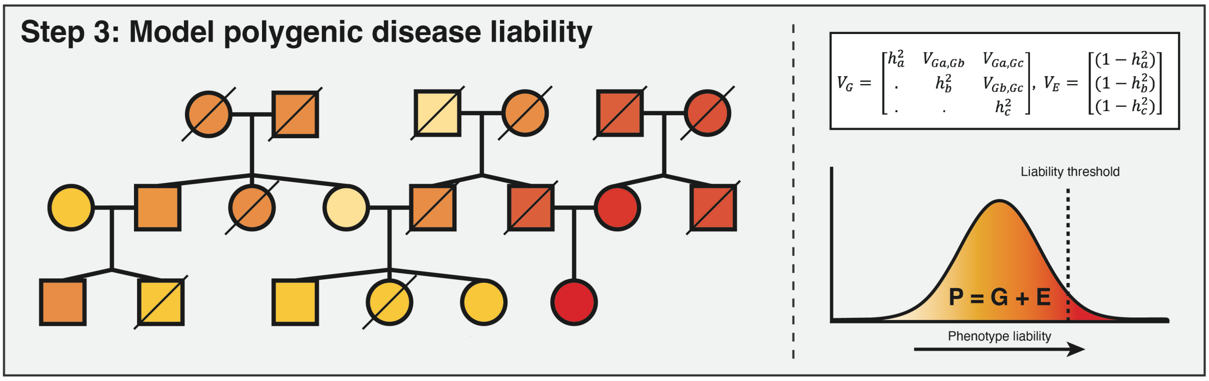


For the polygenic model, we first define the genetic variance-covariance matrix that describes the (polygenic) heritability and co-heritabilities of the three diseases: (a) ALS, (b) FTD and (c) other dementias. Liability was standardised such that *P ~N(0,1)*, ensuring that the liability threshold corresponds directly to the specified lifetime risk.

$$P = G + E, \text{ with } V_G = \begin{bmatrix} h_a^2 & V_{Ga,Gb} & V_{Ga,Gc} \\ . & h_b^2 & V_{Gb,Gc} \\ . & . & h_c^2 \end{bmatrix}, \text{ and } V_E = \begin{bmatrix} (1 - h_a^2) \\ (1 - h_b^2) \\ (1 - h_c^2) \end{bmatrix} \qquad (5)$$

Where $h_a^2$ is the heritability of the first disease (ALS).[8–10] $V_{Ga,Gb}$ is the genetic covariance between disease ALS and FTD defined as:

$$V_{Ga,Gb} = r_{Ga,Gb} * \sqrt{h_a^2 * h_b^2} \qquad (6)$$

With the genetic correlation between the diseases ($r_{Ga,Gb}$) defined in Supplementary Table 1.[11–13] Genetic and non-genetic values in the founder generation and individuals in the external branches for which we have not simulated parents, were drawn from these distributions. Then, for all offspring we draw *G* and *E* from a normal distribution with:

$$G_{offspring} \sim N\left(\frac{G_p + G_m}{2}, \frac{V_G}{2}\right) \qquad (7)$$

Where $G_p$ and $G_m$ are the paternal and maternal genetic values respectively. Note that in the figure we colour by the genetic value for one disease trait, but multiple disease traits are defined simultaneously.

### Step 4: Define disease status

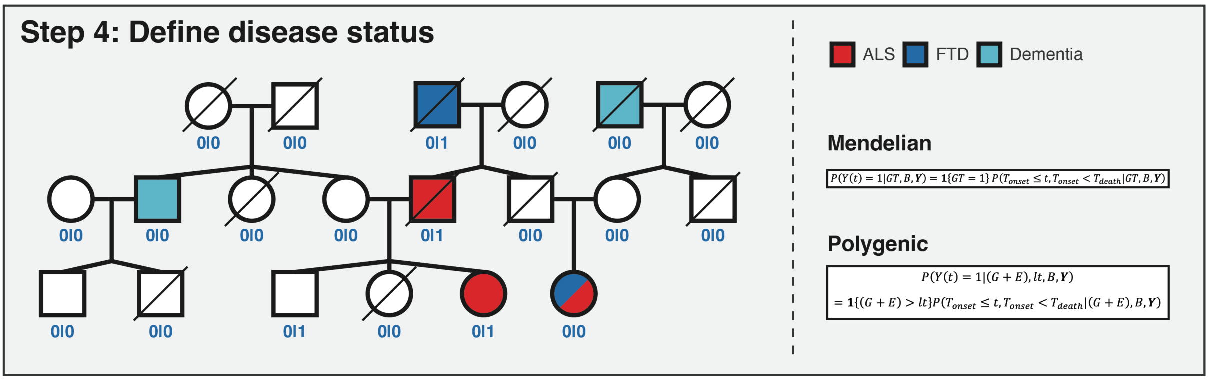


To define disease status, we account for both Mendelian and polygenic disease risk. For monogenic disease, the probability of a carrier being affected is modelled as a Bernoulli draw with probability $p$, where $p$ is the user defined disease penetrance.

$$Affected = \begin{cases} 1 \; if \; carrier \; and \; penetrant \\ 0 \; otherwise \end{cases} \quad (8)$$

For polygenic disease, an individual is affected if the phenotypic liability *P (G+E)* surpasses the liability threshold *lt*. We use the disease lifetime risk *K* to define the liability threshold *lt* such that a proportion *K* of the population exceeds the threshold.[9,14,15] The disease lifetime risk is corrected for the proportion of overall disease attributed to polygenic disease. We thus define *lt* and polygenic disease status as:

$$K_{polygenic} = K_{overall} * (1 - proportion_{mendelian}) \quad (9)$$

$$lt = -\Phi^{-1}(K_{polygenic}) \quad (10)$$

$$Affected = \begin{cases} 1 \; if \; P > lt \\ 0 \; otherwise \end{cases} \quad (11)$$

Final affected status is assigned only if disease onset occurs before censoring, accounting for all-cause mortality and competing diagnoses. We model age at disease onset, based on disease-specific age-at-onset distributions (Figure 3).[5,15–18] We model competing risk from all-cause mortality using an age-specific life expectancy distribution based on the individual's year of birth, as estimated in Step 1 (Figure 1). We model competing risks from other diagnoses using disease specific survival after onset distributions (Figure 4, Figure 5).[19–22]

### Figure 3: Simulation of age at onset

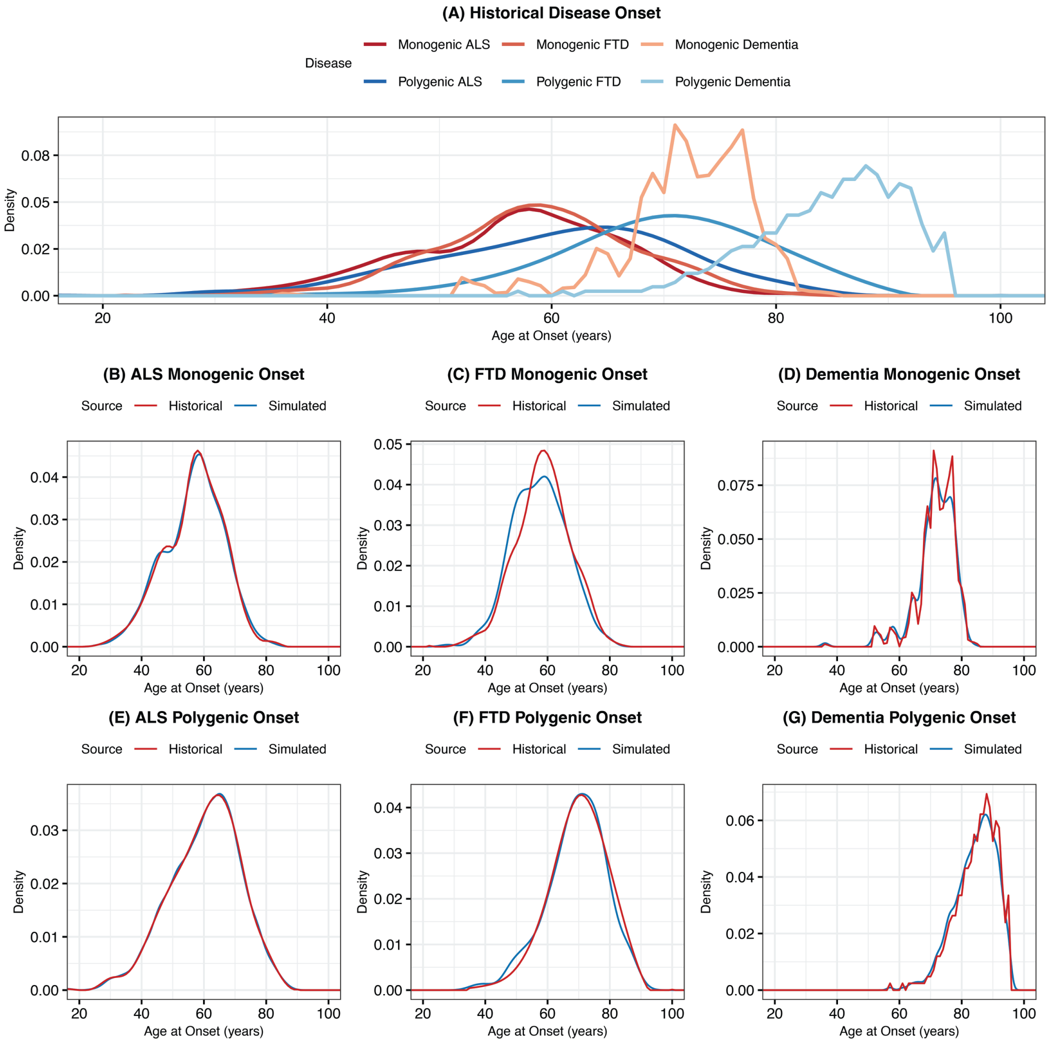

**Figure 4: Simulation of survival after onset**

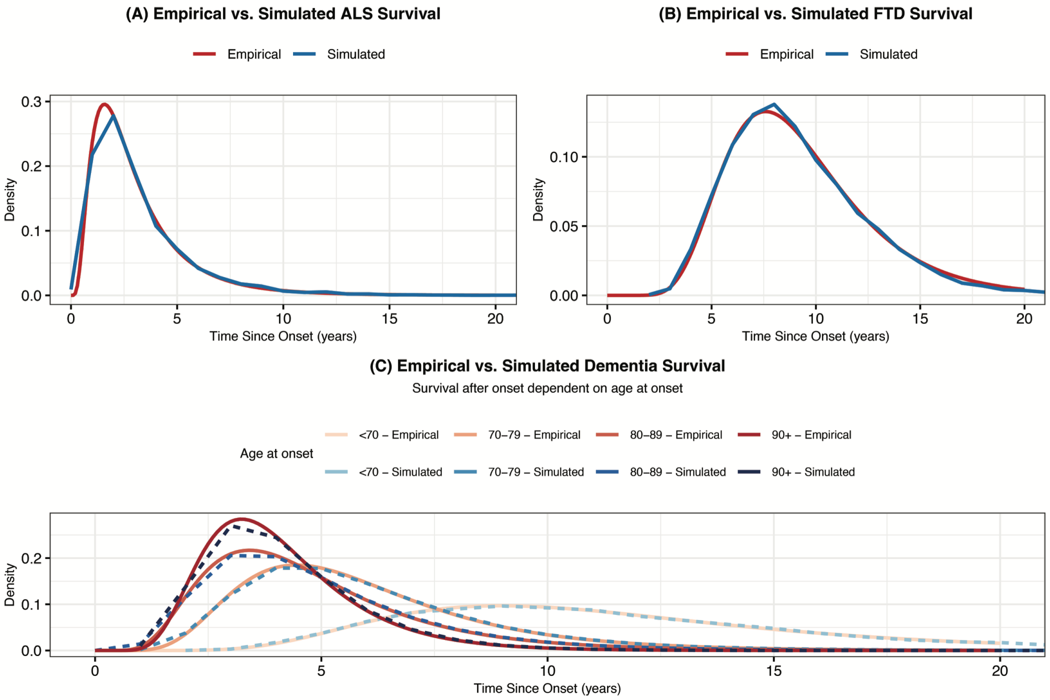


For the Mendelian disease component, the probability of disease by age *t* is defined as:

$$P(Y(t) = 1|GT, B, Y) = 1\{GT = 1\}P(T_{onset} \leq t, T_{onset} < T_{death}|GT, B, Y) \qquad (12)$$

Where *GT* denotes the genotype at the Mendelian disease locus, *B* denotes the individual's year of birth, and *Y* denotes the competing mortality from other diseases. We model age-related penetrance as the probability that disease onset occurs by age *t* and before death from other causes, including competing diagnoses and all-cause mortality.

For the polygenic disease component, the probability of disease by age *t* is defined as

$$P(Y(t) = 1|(G + E), lt, B, Y)$$

$$= 1\{(G + E) > lt\}P(T_{onset} \leq t, T_{onset} < T_{death}|(G + E), B, Y) \qquad (13)$$

Where *G*+*E* denotes the phenotypic liability, *lt* the lifetime risk dependent liability threshold, *B* denotes the individual's year of birth, and *Y* denotes the competing mortality from other diseases. We model age-related polygenic disease onset as the probability that disease onset occurs by age *t* and before death from other causes, including competing diagnoses and all-cause mortality.

**Figure 5: Accounting for competing risk**

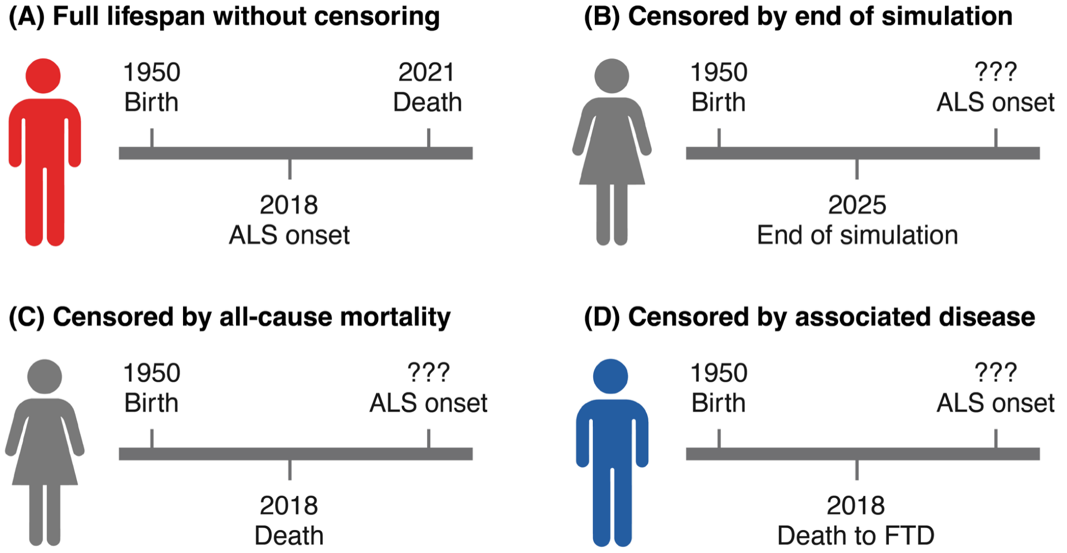


### Step 5: Count affected relatives

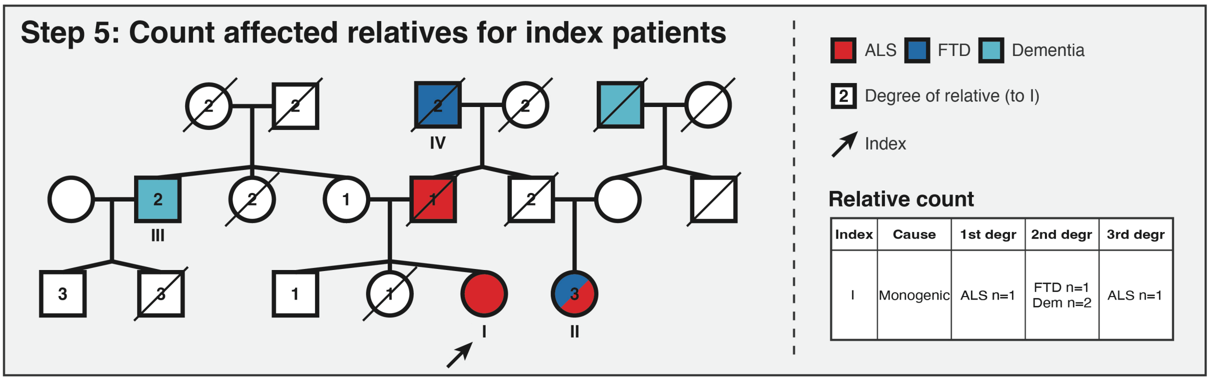


For all ALS index patients with disease onset in the 15 years before the year of simulation, we count the number of affected relatives. The degree of relatives to an index patient is illustrated in the 'Step 5' figure. For relatives up to the third degree, we count relatives with (1) only ALS; (2) ALS and/or FTD; or (3) ALS and/or any dementia. In case of comorbid ALS-FTD or ALS-dementia, we only count unique FTD/dementia cases once (see third-degree individual II in the 'Step 5' figure). For any dementia, we consider both FTD cases and other dementia cases (see second-degree relatives III and IV). We only count affected relatives at the time of disease onset. If a relative of an index would later develop ALS, we do not take that into account for the index patient.

### References Supplementary File 1A

### Supplementary File 1B: Logistic regression models

Logistic regression models were used to assess the association between monogenic ALS status (dependent variable) and family history predictors. Prediction model development proceeded in a stepwise manner. First, we used a model considering ALS-affected relatives, expanding from relatives in the first, to second, to third degree (Step 1). Second, we expanded the prediction model also considering FTD-affected relatives and dementia-affected relatives (Step 2). Finally, we expanded these prediction models by also considering the number of unaffected relatives, instead of just the ALS, ALS and FTD, or ALS and dementia-affected relatives (Step 3).

Step 1: Expanding predictors by degree of relatedness

$$Monogenic\ ALS = \beta_0 + \beta_1(ALS\ first\ degree) \quad (1)$$

$$Monogenic\ ALS = \beta_0 + \beta_1(ALS\ first\ degree) + \beta_2(ALS\ second\ degree) \quad (2)$$

$$Monogenic\ ALS = \beta_0 + \beta_1(ALS\ first\ degree) + \beta_2(ALS\ second\ degree) + \beta_3(ALS\ third\ degree) \quad (3)$$

Step 2: Including relatives with FTD or dementia

$$Monogenic\ ALS = \beta_0 + \beta_1(ALS\ first\ degree) + \beta_2(ALS\ second\ degree) + \beta_3(ALS\ third\ degree) \quad (4)$$

$$Monogenic\ ALS = \beta_0 + \beta_1(ALS\ first\ degree) + \beta_2(ALS\ second\ degree) + \beta_3(ALS\ third\ degree) + \beta_4(FTD\ first\ degree) + \beta_5(FTD\ second\ degree) + \beta_6(FTD\ third\ degree) \quad (5)$$

$$Monogenic\ ALS = \beta_0 + \beta_1(ALS\ first\ degree) + \beta_2(ALS\ second\ degree) + \beta_3(ALS\ third\ degree) + \beta_4(DEM\ first\ degree) + \beta_5(DEM\ second\ degree) + \beta_6(DEM\ third\ degree) \quad (6)$$

Step 3: Including unaffected relatives

Models in Step 3 adjusted for family size by adding unaffected relatives as covariates. In function (7), relatives with FTD were treated as unaffected (ALS outcome only). In function (8), relatives with non-FTD dementia were treated as unaffected (ALS + FTD outcome). In function (9), only relatives without ALS, FTD, or dementia were labelled unaffected.

$$Monogenic\ ALS = \beta_0 + \beta_1(ALS\ first\ degree) + \beta_2(ALS\ second\ degree) + \beta_3(ALS\ third\ degree) + \beta_4(Unaffected\ first\ degree) + \beta_5(Unaffected\ second\ degree) + \beta_6(Unaffected\ third\ degree) \quad (7)$$

$$Monogenic\ ALS = \beta_0 + \beta_1(ALS\ first\ degree) + \beta_2(ALS\ second\ degree) + \beta_3(ALS\ third\ degree) + \beta_4(FTD\ first\ degree) + \beta_5(FTD\ second\ degree) + \beta_6(FTD\ third\ degree) + \beta_7(Unaffected\ first\ degree) + \beta_8(Unaffected\ second\ degree) + \beta_9(Unaffected\ third\ degree) \quad (8)$$

$$Monogenic\ ALS = \beta_0 + \beta_1(ALS\ first\ degree) + \beta_2(ALS\ second\ degree) + \beta_3(ALS\ third\ degree) + \beta_4(DEM\ first\ degree) + \beta_5(DEM\ second\ degree) + \beta_6(DEM\ third\ degree) + \beta_7(Unaffected\ first\ degree) + \beta_8(Unaffected\ second\ degree) + \beta_9(Unaffected\ third\ degree) \quad (9)$$
