## Supplementary Tables for "The value of a family history to distinguish monogenic from polygenic amyotrophic lateral sclerosis"

**Supplementary Table 1: Simulation input parameters and literature estimates**

| Disease parameter | Simulation input | Literature estimates | Sensitivity analyses ranges | Source |
|---|---|---|---|---|
| **Demographic parameters** | | | | |
| Fertility rate | Based on offspring birthyear | Based on offspring birthyear | 1 – 5 children | Gapminder Fertility Rates[7] |
| Life expectancy at age 15 | Based on individual birthyear | Based on individual birthyear | 60 – 100 years | Our World in Data, 2025[11] |
| **Disease allele frequencies** | | | | |
| *ALS-FTD moderate* penetrance allele frequency | 0.075% | 0.078% (*C9orf72*)<br>0.072% (*C9orf72*) | 0.05% – 0.20% | Douglas, 2024[8]<br>Beck, 2013[12] |
| *ALS-FTD high* penetrance allele frequency[a] | 0.012% | 0.017% (*FUS* + *SOD1*)<br>0.012% (*FUS* + *SOD1*) | 0.005% – 0.020% | Douglas 2024[8]<br>gnomAD v4.1.0 |
| *FTD-specific* allele frequency[a] | 0.015% | 0.015% (*GRN* + *MAPT*) | - | gnomAD v4.1.0 |
| **ALS parameters** | | | | |
| *ALS-FTD moderate* allele penetrance | 21% | 21.0% (*C9orf72*)<br>24.1% (*C9orf72*)<br>33.4% (*C9orf72*) | 10 – 50% | Gao, 2025[13]<br>Van Wijk, 2024[9]<br>Douglas, 2024[8] |
| *ALS-FTD high* allele penetrance[b] | 50% | 53.9% (*SOD1*)<br>19.2% (*FUS*) | 30 – 70% | Douglas, 2024[8]<br>Douglas, 2024[8] |
| *FTD-specific* allele penetrance[c] | 0% | 0% (*GRN/MAPT*) | - | Not modelled |
| Lifetime risk[d] | 0.267% * 85%<br>(1 in 375 people) | 0.29% (men)<br>0.23% (women) | 0.22% – 0.33% * 85%<br>(1 in 300-450 people) | Ryan, 2019[14] |
| Heritability | 40% | 43.0%<br>37.0% | 20% – 60% | Trabjerg, 2020[15]<br>Ryan, 2019[14] |
| Monogenic age at onset | Age-specific incidence rates | Age-specific incidence rates | ±10 years | Murphy, 2017[16] |
| Polygenic age at onset | Age-specific incidence rates | Age-specific incidence rates | ±10 years | Nona, 2022[17] |
| **FTD parameters** | | | | |
| *ALS-FTD moderate* allele penetrance | 10% | 10% (*C9orf72*) | - | Gao, 2025[13] |
| *ALS-FTD high* allele penetrance[e] | 10% | - | - | No available source |
| *FTD-specific* allele penetrance | 90% | 90% (*GRN*)<br>>90% (*MAPT*) | - | Van Swieten, 2008[18]<br>Van Swieten, 2008[18] |
| Lifetime risk[d] | 0.134% * 75%<br>(1 in 742 people) | 0.134% (1/742 people) | - | Coyle-Gilchrist, 2016[19] |
| Heritability | 45% | 48% | - | Dijkstra, 2025[20] |
| Monogenic age at onset | Age-specific incidence rates | Age-specific incidence rates | - | Murphy, 2017[16] |
| Polygenic age at onset | Age-specific incidence rates | Age-specific incidence rates | - | Nilsson, 2014[21] |
| **Dementia parameters** | | | | |
| *ALS-FTD moderate* allele penetrance[f] | 50% | 59% (*C9orf72*) | - | Gao, 2025[13] |
| *ALS-FTD high* allele penetrance[e] | 10% | - | - | No available source |
| *FTD-specific* allele penetrance[c] | 0% | - | - | Not modelled |
| Lifetime risk | 41.8% | 41.8% | - | Fang, 2025[22] |
| Heritability[g] | 55% | 73% (Alzheimer's disease) | - | Dijkstra, 2025[20] |
| Monogenic age at onset | Age-specific incidence rates, competing mortality | Age-specific incidence rates, competing mortality | - | Gao, 2025[13] |
| Polygenic age at onset | Age-specific incidence rates, competing mortality | Age-specific incidence rates, competing mortality | - | Fang, 2025[22] |
| **Genetic correlations** | | | | |
| ALS ~ FTD | 60% | 58.6%<br>63.7% | 40% – 80% | Van Rheenen, 2021[23]<br>Chen, 2024[24] |

| | | | | |
|---|---|---|---|---|
| ALS ~ Dementia | 25% | 31.2%<br>27.3%<br>16.0%-25.0% | - | Van Rheenen, 2021[23]<br>Chen, 2024[24]<br>Wainberg, 2023[25] |
| FTD ~ Dementia | 35% | 22.9%<br>38.6% | - | Van Rheenen, 2021[23]<br>Chen, 2024[24] |
| **Survival after disease onset** | | | | |
| ALS (median years, IQR) | 2.5 (1.7 – 4.0) | 2.5 (1.7 – 4.0)<br>2.2 (1.1 – 4.4) | - | Engelberg-Cook, 2024[26]<br>Pupillo, 2025[27] |
| FTD (median years, IQR) | 8.7 (6.7 – 11.8) | 8.7 (6.7 – 11.8) | - | Foxe, 2025[28] |
| Dementia[h] (median years, IQR) | Age of onset dependent | 6.5 (4.8 – 8.5)<br>4.5 (2.8 – 7.0) | - | Foxe, 2025[28]<br>Xie, 2008[29] |

[a]Allele frequency represents the compound allele frequencies for pathogenic *FUS+SOD1* and *GRN+MAPT* mutations.

[b]Combined penetrance of *FUS* and *SOD1* was applied for the compound '*ALS-FTD high* penetrance' disease allele.

[c]No ALS phenotype was modelled for carriers of the *FTD-specific* disease allele. Given the high penetrance for FTD of the *FTD-specific* allele, no additional 'other dementia' was simulated.

[d]Lifetime risk was multiplied by the percentage of disease accounted for by polygenic disease, as literature estimates of lifetime risks account for overall (monogenic and polygenic) disease.

[e]No reported penetrance for FTD or 'any dementia' associated with *FUS/SOD1* mutations was available. FTD penetrance was set at 10% to account for FTD associated with non-*C9orf72*, rare, high-penetrance ALS-associated mutations.

[f]Dementia penetrance of the '*ALS-FTD moderate* penetrance' disease allele was lowered in simulations, as Gao *et al.*[13] calculated 'any dementia' penetrance including 10% FTD penetrance, which was already modelled under FTD specific parameters.

[g]Heritability was lowered compared to Dijkstra *et al.*[20] to account for any form of dementia. We assumed lower heritability for e.g. vascular dementias.

[h]Survival dependent on age at onset with earlier onset correlating to longer survival.

**Supplementary Table 2: Varying input parameters for sensitivity analyses**

| Input parameter | Main simulation input | Sensitivity analysis input | Sensitivity range | Positive predictive value range |
|---|---|---|---|---|
| Fertility rate[a] | Dutch historical data | 1 – 5 children | 20.0% – 56.4% | 86.8% – 93.9% |
| Life expectancy[a] | Dutch historical data | 60 – 100 years | 25.8% – 50.2% | 84.7% – 94.6% |
| Age at onset monogenic ALS[b] | Published *C9orf72*-ALS age at onset distribution | ±10 years | 33.9% – 44.3% | 79.9% – 89.2% |
| Age at onset polygenic ALS[b] | Published ALS age at onset distribution | ±10 years | 40.7% – 44.3% | 87.9% – 93.2% |
| *ALS-FTD moderate* allele frequency | 0.075% | 0.05% – 0.20% | 38.3% – 44.0% | 85.3% – 93.5% |
| *ALS-FTD moderate* allele penetrance | 21% | 10 – 50% | 40.3% – 64.7% | 82.9% – 95.6% |
| *ALS-FTD high* allele frequency | 0.012% | 0.005% – 0.020% | 37.9% – 49.5% | 85.6% – 91.0% |
| *ALS-FTD high* allele penetrance | 50% | 30 – 70% | 33.6% – 50.1% | 83.5% – 90.7% |
| ALS lifetime risk[c] | 0.267% * 85% (1 in 375) | 0.22% – 0.33% * 85% (1 in 300-450) | 38.6% – 43.0% | 81.4% – 93.1% |
| ALS heritability | 40% | 20% – 60% | 37.2% – 45.1% | 78.8% – 93.5% |
| Genetic correlation ALS ~ FTD | 60% | 40% – 80% | 40.7% – 42.9% | 87.3% – 88.8% |
| Censoring year[d] | 2025 | 1985 – 2065 | 34.5% – 48.3% | 85.9% – 91.3% |

[a]Mean fertility rate and life expectancy trends vary over time. Sensitivity analyses were performed with a set value for all individuals, regardless of year of birth.

[b]Age at onset for monogenic and polygenic disease was derived from prior published studies. Sensitivity analyses then shifted distributions ±10 years to simulate the effect of monogenic/polygenic ALS occurring at earlier/later age.

[c]Overall ALS lifetime risk was corrected for the percentage of disease attributable to polygenic ALS, as the lifetime risk was used as a disease parameter for polygenic disease.

[d]Censoring year is the year in which the simulation is run. For each censoring year, potential index patients birthyears are sampled from 25 to 85 years prior to the censoring year. Only patients with disease onset in the 15 years before censoring are included for fALS criteria accuracy assessment.

**Supplementary Table 3: Accuracy comparison between traditional fALS criteria and logistic prediction models**

| fALS criteria / model | Sensitivity | Positive predictive value | False positive rate |
|---|---|---|---|
| **Relatives with ALS** | | | |
| *fALS criteria:* ≥2 relatives within second degree | 31.1% | 90.0% | 10.0% |
| *Prediction model:* Considering relatives within third degree | 31.5% | 91.2% | 8.8% |
| **Relatives with ALS or FTD** | | | |
| *fALS criteria:* ≥2 relatives within second degree | 43.3% | 90.2% | 9.8% |
| *Prediction model:* Considering relatives within third degree | 43.9% | 91.8% | 8.2% |
| **Relatives with ALS or any dementia** | | | |
| *fALS criteria:* ≥2 relatives within second degree | 63.5% | 64.1% | 35.9% |
| *Prediction model:* Considering relatives within third degree | 63.6% | 75.1% | 24.9% |

Comparison of predictive accuracy between traditional fALS criteria and logistic prediction models, predicting underlying monogenic inheritance based on number of affected and unaffected relatives.

**Supplementary Table 4: fALS criteria cutoffs with high positive predictive value for monogenic disease**

| fALS criteria | Percentage of patients[a] | Sensitivity[b] | Positive predictive value[b] | False positive rate[b] |
|---|---|---|---|---|
| **≥3 affected relatives** | | | | |
| 1st-2nd degree with ALS or FTD | 3.1% | 18.4% (16.0%-21.0%) | 95.9% (92.0%-97.8%) | 4.1% (2.2%-8.0%) |
| 1st-3rd degree with ALS or FTD | 5.1% | 28.7% (25.8%-31.7%) | 93.1% (89.5%-95.5%) | 6.9% (4.5%-10.5%) |
| 1st-3rd degree with ALS | 3.5% | 20.0% (17.6%-22.8%) | 94.2% (90.1%-96.6%) | 5.8% (3.4%-9.9%) |
| **≥4 affected relatives** | | | | |
| 1st-2nd degree with ALS or FTD | 1.2% | 6.8% (5.4%-8.7%) | 96.8% (90.0%-98.2%) | 3.2% (1.8%-10.0%) |
| 1st-3rd degree with ALS or FTD | 2.7% | 15.6% (13.3%-18.1%) | 95.2% (90.7%-97.4%) | 4.8% (2.6%-9.3%) |
| 1st-3rd degree with ALS | 1.8% | 10.8% (8.9%-12.9%) | 97.0% (92.0%-98.5%) | 3.0% (1.5%-8.0%) |

Predictive accuracy of various fALS definitions predicting monogenic inheritance based on number of affected relatives of an ALS-affected index patient in three-generation pedigrees.
[a]Percentage of index cases (n=5,431) that matched fALS criterion.
[b]Values shown as: predictive measure percentage (95% Confidence Intervals).
